# Robust, credible, and interpretable AI-based histopathological prostate cancer grading

**DOI:** 10.1101/2024.07.09.24310082

**Authors:** Fabian Westhaeusser, Patrick Fuhlert, Esther Dietrich, Maximilian Lennartz, Robin Khatri, Nico Kaiser, Pontus Röbeck, Roman Bülow, Saskia von Stillfried, Anja Witte, Sam Ladjevardi, Anders Drotte, Peter Severgardh, Jan Baumbach, Victor G. Puelles, Michael Häggman, Michael Brehler, Peter Boor, Peter Walhagen, Anca Dragomir, Christer Busch, Markus Graefen, Ewert Bengtsson, Guido Sauter, Marina Zimmermann, Stefan Bonn

## Abstract

**Background:** Prostate cancer (PCa) is among the most common cancers in men and its diagnosis requires the histopathological evaluation of biopsies by human experts. While several recent artificial intelligence-based (AI) approaches have reached human expert-level PCa grading, they often display significantly reduced performance on external datasets. This reduced performance can be caused by variations in sample preparation, for instance the staining protocol, section thickness, or scanner used. Another limiting factor of contemporary AI-based PCa grading is the prediction of ISUP grades, which leads to the perpetuation of human annotation errors.

**Methods:** We developed the <u>p</u>rostate <u>c</u>ancer <u>a</u>ggressiveness index (PCAI), an AI-based PCa detection and grading framework that is trained on objective patient outcome, rather than subjective ISUP grades. We designed PCAI as a clinical application, containing algorithmic modules that offer robustness to data variation, medical interpretability, and a measure of prediction confidence. To train and evaluate PCAI, we generated a multicentric, retrospective, observational trial consisting of six cohorts with 25,591 patients, 83,864 images, and 5 years of median follow-up from 5 different centers and 3 countries. This includes a high-variance dataset of 8,157 patients and 28,236 images with variations in sample thickness, staining protocol, and scanner, allowing for the systematic evaluation and optimization of model robustness to data variation. The performance of PCAI was assessed on three external test cohorts from two countries, comprising 2,255 patients and 9,437 images.

**Findings:** Using our high-variance datasets, we show how differences in sample processing, particularly slide thickness and staining time, significantly reduce the performance of AI-based PCa grading by up to 6.2 percentage points in the concordance index (C-index). We show how a select set of algorithmic improvements, including domain adversarial training, conferred robustness to data variation, interpretability, and a measure of credibility to PCAI. These changes lead to significant prediction improvement across two biopsy cohorts and one TMA cohort, systematically exceeding expert ISUP grading in C-index and AUROC by up to 22 percentage points.

**Interpretation:** Data variation poses serious risks for AI-based histopathological PCa grading, even when models are trained on large datasets. Algorithmic improvements for model robustness, interpretability, credibility, and training on high-variance data as well as outcome-based severity prediction gives rise to robust models with above ISUP-level PCa grading performance.

## Introduction

Prostate cancer (PCa) stands as a significant public health concern worldwide, representing a challenge in both clinical management and public health care systems. PCa ranks among the most prevalent cancers in men, with approximately 1.4 million new cases worldwide each year with its incidence steadily increasing over the past decades ^1^. Due to the wide variety in growth rates of PCa, histopathology plays a central role in the diagnosis and management of PCa. An essential part of PCa diagnosis is the histopathological assessment of prostate biopsies. The Gleason score is the most relevant prognostic feature in prostate cancer biopsies^2^ based on Gleason grading ^3^. The International Society of Urological Pathology (ISUP) has proposed a simplified system defining ISUP groups 1-5 with increasing cancer severity to predict disease aggressiveness ^4^. For biopsy assessment, the most frequent and worst Gleason grades are combined to form a Gleason score (e.g. 3+4). ISUP then rearranges the Gleason scores into five ISUP grades, assigning the groups 1 (3+3 and below), 2 (3+4), 3 (4+3), 4 (3+5, 4+4, 5+3) and 5 (4+5, 5+4, 5+5). These categories are used to guide the urologist in treatment decisions. Unfortunately, even between expert pathologists the concordance in Gleason grading suffers from high interobserver variability, leading to possible over- or under-treatment due to the subjective nature of the visual assessment ^5^.

Recent advancements in digital pathology, including the introduction of high throughput digital slide scanners, hold the potential to improve histopathological evaluation of PCa. The possibility to use algorithms to automatically analyze pathological samples is not only time and cost effective but offers the potential for a standardized, objective, and accurate evaluation, providing crucial insights into tumor characteristics, aiding in treatment decisions, and assessing disease progression. This offers the opportunity to an efficient and reproducible histopathological assessment, thereby optimizing diagnostic accuracy and streamlining workflows in PCa diagnosis and care.

Many automated PCa grading approaches are based on the traditional Gleason or ISUP score. Several of these algorithms have attained pathologist-level performance in these tasks ^6–8^. Model training on the ISUP score, however, is limited in its usefulness, as it distinguishes only five subgroups, one of which is characterized by a very good (ISUP 1) and three of which are characterized by a bad prognosis (ISUP 3-5). Moreover, models trained on human ISUP annotations will replicate human error, which is why recent approaches have shifted towards the more interpretable prediction of objective endpoints, such as time to relapse (e.g. biochemical recurrence or cancer-related death). Some of these studies concentrate on modeling the probability of relapse-free survival over time ^9^, while others predict the probabilities of relapse up to one or multiple fixed time points ^10^. While many promising steps have been conducted in the automated assessment of PCa histopathological data, four prominent challenges still require further attention: Robustness, interpretability, trustworthiness, and above ISUP-level grading performance ^11^.

The biggest hurdle for automated PCa grading, possibly, is the variation of histopathological protocols, which can result in a lack of robustness of algorithms ^11–13^. Processing tissue for digitization consists of several steps: tissue formalin fixation, paraffin embedding, sectioning, staining and digitization with a slide scanner. Each step involves numerous parameters that can vary between clinics, research institutions and even within the same lab leading to variations in the appearance of the tissue in the images. Especially AI-based PCa grading seems to be negatively affected by data variation ^11^, while PCa detection seems to be robust to data variation and is already in clinical use ^14,15^. Given the potential data variation-induced degradation of the predictive performance of AI-based PCa, concepts of model trustworthiness have recently been proposed ^16,17^. Model trustworthiness allows for the deferral of PCa grading of problematic samples to a human expert. Of note, a systematic investigation of the impact of data variation on automated PCa grading, as for example conducted for breast cancer ^17^, is still missing.

In this work, we first systematically assess the impact of data variation on AI-based PCa grading and subsequently show how algorithmic improvements can result in robust, interpretable, and credible model predictions with state-of-the-art performance. To this end, we performed a multicentric, retrospective, observational trial that contains six cohorts with over 25,591 patients, 83,864 images, and 5 years of median follow-up from 5 different centers and 3 countries. An important part of this data is a unique high-variance cohort of 8,157 patients with 28,236 scanned TMAs with variations in sample thickness, staining time, and scanner. The data also contains patient relapse information for 5 years on average, which our algorithms use as an objective measure for cancer aggressiveness, instead of replicating the subjective Gleason score. Using this dataset, we systematically evaluated the robustness of AI-based PCa grading to data variation. We observed severe performance degradation for e.g. variations in section thickness and staining time with our baseline model. The addition of a select set of algorithmic improvements to the baseline model created PCAI, which is robust to data variation, interpretable, and contains a measure of credibility. These changes lead to significant prediction improvement across all evaluated datasets, systematically outperforming the five tier Gleason (ISUP) grading in both the Concordance Index (C-Index) and area under the curve (AUROC) across three different external test cohorts.

## Methods

### Data acquisition and composition

The primary aim of this work is to build an algorithm that is robust, explainable, and credible and exceeds human PCa grading performance, which necessitates a large, heterogeneous dataset with rich patient follow-up information of sufficient quality.

The standard for clinical diagnosis is ISUP grading of preoperative biopsies, typically obtained through transrectal ultrasound-guided biopsy. Multiple tissue samples are collected from different areas of the prostate gland to improve cancer detection rates. After biopsy, specimens are formalin-fixed and paraffin-embedded to preserve structure and stained with Haematoxylin and Eosin (H&E) to enhance cellular visibility for pathologist examination. All biopsies in this work contain ISUP grades by expert pathologists. Biopsies have long edge lengths in the order of 60,000 pixels with a total of up to 10 billion pixels per image.

In addition to biopsies, this work uses postoperative tissue microarrays (TMAs) from radical prostatectomies. TMAs consist of many small cylindrical representative samples, termed spots, that are extracted from paraffin-embedded tissue and are widely used in biomarker discovery and validation studies. TMA spots of this work typically have edge lengths of 3,000 to 6,000 pixels with resulting images that contain in the order of 10 million pixels and are much smaller than biopsies. TMA spots are preselected to represent a patient’s cancer status. All ISUP grade annotations for TMA spots were retrieved from the routine diagnostic reports made by expert pathologists’ examinations of the resected whole prostate to derive a patient-level annotation. Therefore, TMA spots might only partially capture a patient’s cancer morphology, with the notable exception of the UKE-sealed dataset, which contains TMA spot-based ISUP grades (see UKE-sealed section).

To the best of our knowledge, we collected the biggest and most heterogeneous histopathological PCa datasets to date, with a total of 81,572 TMA spot images and 3,388 biopsy images retrospectively collected from 25,591 patients of five different clinics with follow-up information of up to 23 years and a maximum of 8 images for a single patient (Fig. 1). This dataset is divided into several subsets acquired with different parameters and used for training or testing the model on images with high variation. Detailed information on demographics and metadata distributions can be found in Table 1. Datasets that were used to build and assess robustness of the proposed model are shown in Fig. 1B. All image-level annotated, unseen datasets that were exclusively used for evaluation are depicted in Fig. 1C. The largest subset is the UKE high variance cohort provided by the University Medical Center Hamburg Eppendorf which contains 17,700 patients who underwent radical prostatectomy (RP) with a follow-up time up to 23 years. This unique dataset allows us to assess differences in acquisition protocol parameters and represents the foundation for building a robust prediction model in this work.

**Figure 1.**
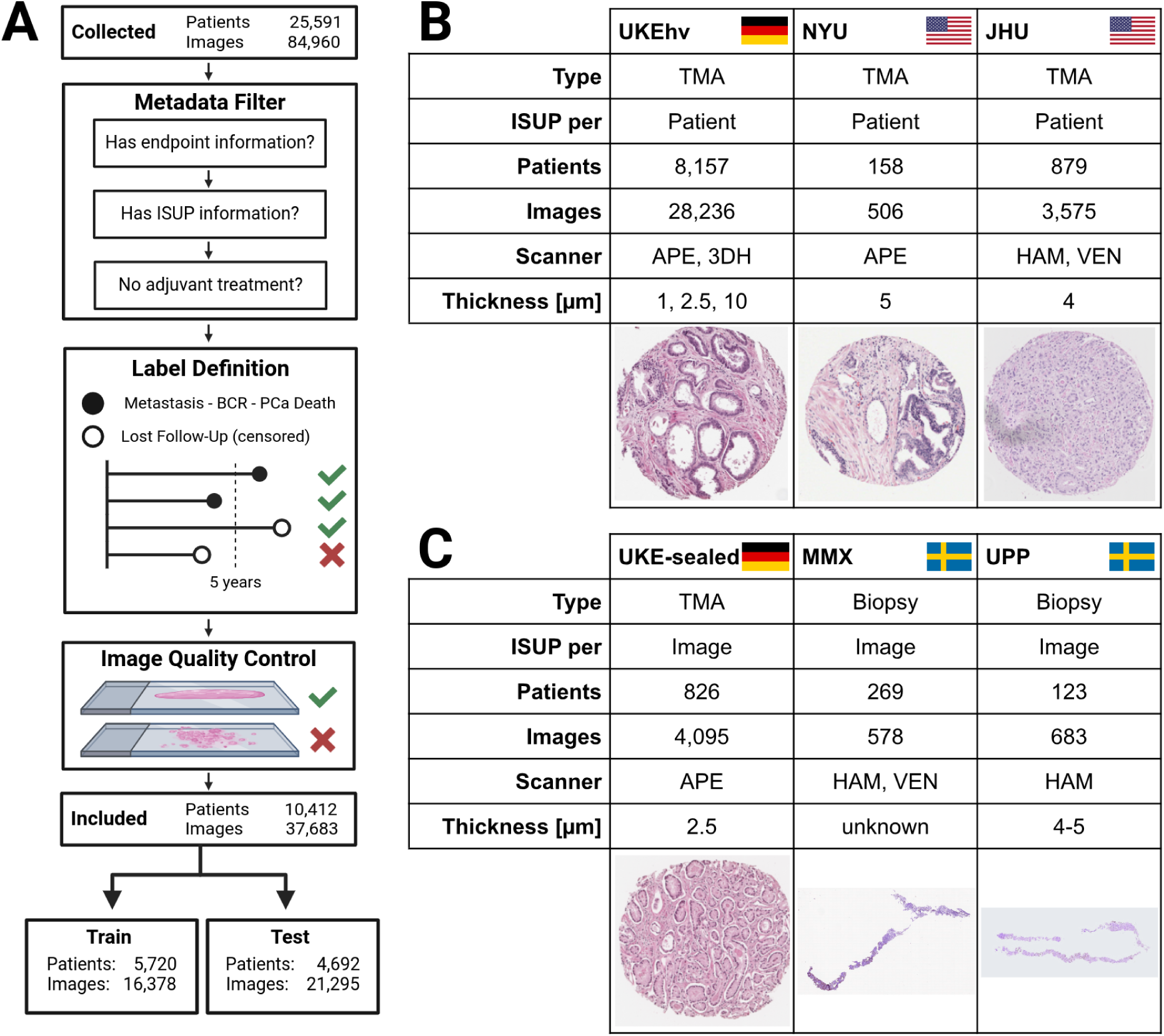
Overview of data preprocessing and the TMA and biopsy datasets. In total, data from 5 different clinics across three countries were collected and integrated. Filtering was performed on the individual patient’s metadata (sufficient endpoint information, minimum 5 years of follow-up (FU) duration or any observed event, ISUP information) and image quality (enough tissue on slide, slide not blurry). **A** shows the preprocessing and filtering of all datasets that are divided into training and test sets. **B** depicts the datasets used to build and assess the robustness of the deep learning algorithm. From the UKEhv dataset, which includes highly variant data from six different domains, three are used for training PCAI. The remaining three domains as well as the external JHU and NYU dataset are used to evaluate robust performance on unseen data that expresses a covariate shift. **C** depicts the data used for benchmarking our algorithm against human annotated IQ Gleason and ISUP on a per-image level. Predictive performance is assessed on one unseen TMA-spot (UKE-sealed) and two external biopsy datasets (MMX, UPP). Scanner vendors: APE=Aperio, 3DH=3DHistech, HAM=Hamamatsu, VEN=Ventana.

**Table 1.** Basic patient characteristics of all experiments showing number of unique patients, number of images and image type, age, PSA level at diagnosis (NYU, JHU), biopsy (MMX) or RP (Other), censoring rate, median survival and follow-up time in months, the event type classification (BCR=biochemical recurrence, META=metastasis, TRT=any additional treatment, FU=lost to follow-up, PCAD=PCa death), primary and secondary Gleason score, ISUP, pathological (TMA), and clinical (biopsy) T-, N- and M-stage.

|  |  | UKE | NYU | JHU | UPP | MMX |
| --- | --- | --- | --- | --- | --- | --- |
|  | patients | 8157 | 158 | 879 | 123 | 269 |
|  | image type | TMA | TMA | TMA | Biopsy | Biopsy |
| | age [years], mean ± SD | 63.5 ± 6.1 | 60.9 ± 7 | 59.2 ± 6.3 | | 67.6 ± 8.9 |
|  | censoring [%] | 61.4 | 70.3 | 0.3 | 83.7 | 88.5 |
|  | median survival [years] | 1.6 | 3.9 | 2 | 2.1 | 4.3 |
|  | median followup [years] | 8 | 17.8 | 16 | 7 | 9.1 |
| ISUP | 0 | 410 (5.03%) |  |  |  |  |
|  | 1 | 1806 (22.14%) | 49 (31.01%) | 133 (15.13%) |  | 15 (5.58%) |
|  | 2 | 4016 (49.23%) | 67 (42.41%) | 337 (38.34%) | 66 (57.89%) | 82 (30.48%) |
|  | 3 | 1367 (16.76%) | 16 (10.13%) | 184 (20.93%) | 27 (23.68%) | 80 (29.74%) |
|  | 4 | 109 (1.34%) | 11 (6.96%) | 123 (13.99%) | 12 (10.53%) | 36 (13.38%) |
|  | 5 | 449 (5.50%) | 15 (9.49%) | 102 (11.60%) | 9 (7.89%) | 56 (20.82%) |
| Gleason score | ≤ 3+3 | 2216 (27.17%) | 49 (31.01%) | 134 (15.28%) | | 15 (5.58%) |
|  | 3+4 | 4016 (49.23%) | 67 (42.41%) | 334 (38.08%) | 66 (57.89%) | 82 (30.48%) |
|  | 3+5 | 37 (0.45%) | 7 (4.43%) | 28 (3.19%) | 1 (0.88%) | 10 (3.72%) |
|  | 4+3 | 1367 (16.76%) | 16 (10.13%) | 185 (21.09%) | 27 (23.68%) | 80 (29.74%) |
|  | 4+4 | 55 (0.67%) | 3 (1.90%) | 84 (9.58%) | 11 (9.65%) | 26 (9.67%) |
|  | 4+5 | 366 (4.49%) | 10 (6.33%) | 85 (9.69%) | 8 (7.02%) | 46 (17.10%) |
|  | 5+3 | 17 (0.21%) | 1 (0.63%) | 10 (1.14%) |  |  |
|  | 5+4 | 82 (1.01%) | 5 (3.16%) | 15 (1.71%) | 1 (0.88%) | 8 (2.97%) |
|  | 5+5 | 1 (0.01%) |  | 2 (0.23%) |  | 2 (0.74%) |
| event type | BCR | 3089 (37.87%) | 43 (27.22%) | 521 (59.27%) | 16 (14.04%) |  |
|  | FU | 5007 (61.38%) | 111 (70.25%) | 3 (0.34%) | 96 (84.21%) | 226 (84.01%) |
|  | META | 61 (0.75%) |  | 142 (16.15%) | 2 (1.75%) | 42 (15.61%) |
|  | PCAD |  | 4 (2.53%) |  |  | 1 (0.37%) |
|  | TRT |  |  | 213 (24.23%) |  |  |
| T-stage | ≤ T1 | 2 (0.02%) | | | 87 (77.68%) | 122 (45.35%) |
|  | T2 | 4966 (60.88%) | 104 (65.82%) | 134 (15.42%) | 25 (22.32%) | 90 (33.46%) |
|  | T3 | 3128 (38.35%) | 52 (32.91%) | 735 (84.58%) |  | 54 (20.07%) |
|  | T4 | 61 (0.75%) | 2 (1.27%) |  |  | 3 (1.12%) |
| N-stage | N0 | 4306 (86.41%) | 56 (35.44%) | 700 (80.18%) |  |  |
|  | N1 | 677 (13.59%) | 1 (0.63%) | 163 (18.67%) |  |  |
|  | N2 |  |  | 2 (0.23%) |  |  |
|  | NX |  | 101 (63.92%) | 8 (0.92%) |  |  |
| M-stage | M0 | 6335 (78.47%) |  | 509 (60.89%) | 7 (6.14%) | 79 (29.48%) |
|  | M1 | 1738 (21.53%) |  | 327 (39.11%) | 6 (5.26%) |  |
|  | MX |  |  |  | 101 (88.60%) | 189 (70.52%) |

As a quantifiable measure correlating with cancer aggressiveness that does not rely on subjective human annotations, we combine the earliest reported indication of disease progression. Possible cancer aggressiveness-related events are biochemical recurrence (BCR), developing metastasis, and PCa-related death with corresponding event times relative to the date of RP for TMA spots or to the date of the biopsy. If none of those exist, the follow-up time is used as the censoring time.

We applied the same metadata and image quality-based filtering steps to all datasets. In brief, we limited patient inclusion to patients that experienced BCR or any other indicator of disease progression (metastasis or PCa-related death) or had at least 5 years of follow-up data available. All patients with a documented adjuvant treatment were removed. Additionally, we excluded images with insufficient quality (e.g. too blurry or no full TMA spot on the image) from the analyses, as summarized in Fig. 1A.

All datasets used in this study were collected in strict accordance with ethical guidelines and compliance regulations. Data collection was approved by the relevant institutional review boards or ethics committees. Informed consent was obtained from all participants involved in data collection processes or the need for informed consent was waived by the local ethics review board. Additionally, any information pertaining to participants was anonymized or de-identified prior to analysis. We will now describe the datasets used in this study in detail.

### UKE-high-variance (UKEhv) TMAs

The UKEhv cohort provided by the University Medical Center Hamburg Eppendorf contains patients who underwent RP between 1992 and 2014 aged 63.8 ± 6.4 years at the UKE with a FU time up to 23 years. The cohort’s observed median PSA level at the time of RP is 6.9 ng/mL (IQR of 4.8 to 10.5 ng/mL). In total, 17,700 patient samples were collected in the TMA dataset, providing 69,251 images. Patients received an annual follow-up ^2^. PSA values were measured following surgery and PSA recurrence was defined as a postoperative PSA of 0.2 ng/mL and increasing at subsequent measurements. Patients without any of these events are considered censored at the last follow-up date. Further, this dataset includes some patients with healthy tissue who therefore did not obtain an ISUP grading.

Building upon this rich information of 17,700 patients, a large variety of 69,251 high-quality images and spots were obtained from different protocols, which represent the foundation for building a robust prediction model in this work. ISUP grades were assigned by examining the whole prostate after RP for every individual patient. After filtering according to the aforementioned criteria, we include 8,157 unique patients and 28,236 TMA spot images as shown in Fig. 1B. This extracted dataset consists of images with varying attributes, like multiple spots for the same patient, varying scanners, section thicknesses and staining times and is, to our knowledge, the largest and most variant collection of TMA spot image data paired with rich follow-up data collected to date. We divided the data into six sub-datasets as depicted in Fig. 2 that were used for training (UKE-first, UKE-second, and UKE-scanner) and testing (all UKEhv datasets). The sub-datasets are described in more detail in the following. Note that all sub-datasets stem from the same patient population and a single patient can contribute images to multiple sub-datasets. Detailed patient-level information for the UKEhv sub-datasets can be found in Table S1.

**Figure 2.**
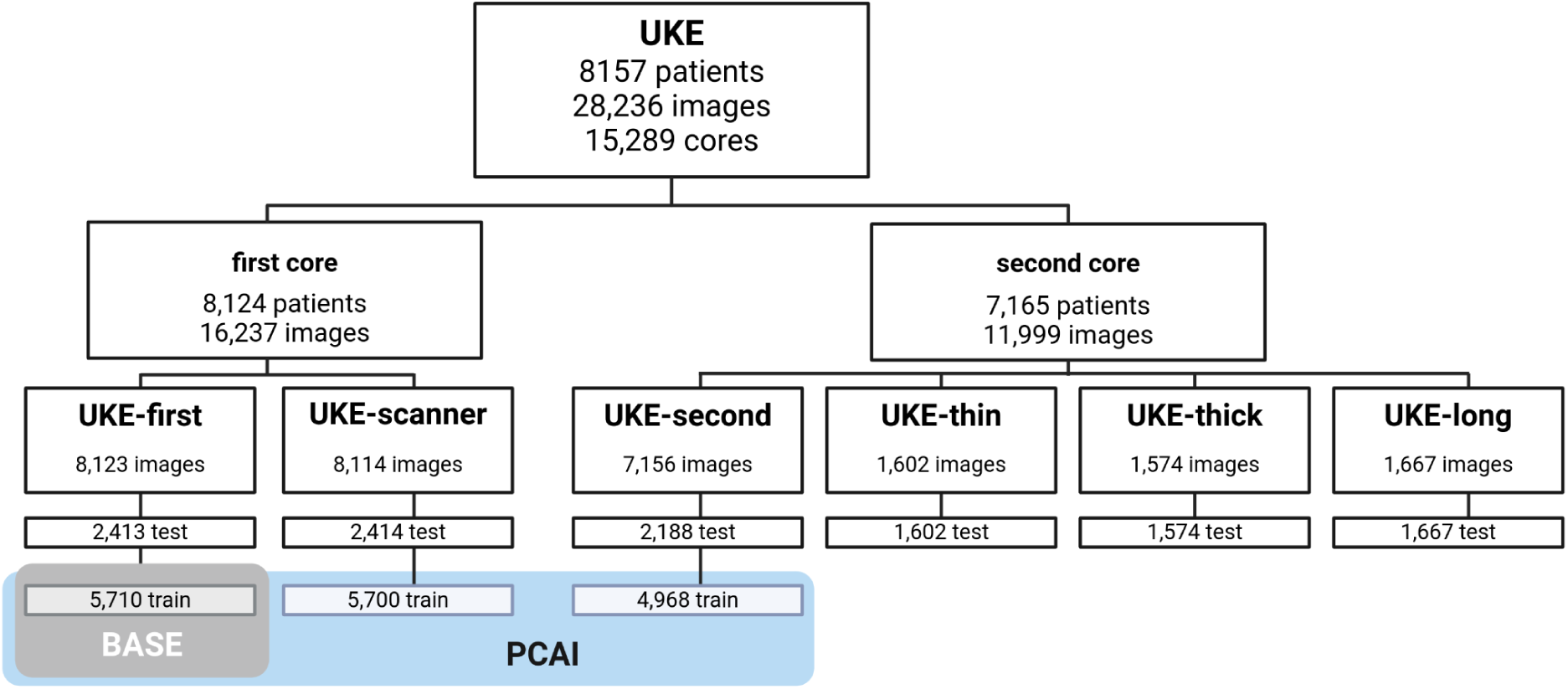
Number of patients and images of the UKEhv sub-datasets per train and test split. The BASE model is trained on TMAs of UKE-first and PCAI on UKE-first, -scanner, and UKE-second training datasets.

### UKE-first

This sub-dataset, used for training and testing of the BASE and PCAI models (see section ‘PCAI design rationale’), encompasses 8,123 tissue TMA spots, each selected to represent the most characteristic spot for ISUP grading within each patient. These spots were utilized both as training and testing data. Patient-level ISUP scores were obtained as part of routine diagnostics. The protocol for digitization followed the standard procedure of the University Medical Center Hamburg Eppendorf (UKE), where tissue samples were sectioned at a thickness of 2.5µm, stained with Hematoxylin and Eosin for 4 minutes and 1:20 minutes, respectively, and then digitized using an Aperio scanner at a magnification of 40x (0.25µm per pixel). Patient event labels were determined by combining biochemical recurrence (BCR), metastasis, or prostate cancer-related death, with patients without any of these events being censored at the last follow-up date.

### UKE-scanner

In this sub-dataset, used for training of PCAI and testing of the BASE and PCAI models, TMA images underwent scanning using a 3DHistech scanner. These images were employed both as training and testing data, with ISUP scores retrieved from routine diagnostics. Following the standard digitization protocol of the UKE, the sub-dataset contains 8,114 images scanned at 80x magnification (0.125µm per pixel).

### UKE-second

Each of the 7,156 images in the UKE-second sub-dataset, used for training of PCAI and testing of the BASE and PCAI models, represents a secondary batch of TMA spots from the cancerous area of the prostate. These TMAs were processed at a different time and underwent slight variations in the protocol. The ISUP scores were retrieved from routine diagnostics, and the digitization protocol followed the standard procedure of the UKE.

### UKE-thin

The UKE-thin sub-dataset, used for testing of the BASE and PCAI models, comprises 1,602 images, each representing a different TMA spot from the cancerous area of the prostate for every patient. These images were exclusively used as testing data. ISUP scores were determined as part of routine diagnostics. Tissue samples were sectioned at 1µm thickness, following the standard digitization protocol of the UKE.

### UKE-thick

The UKE-thick sub-dataset, comprising 1,574 images and used for testing of the BASE and PCAI models, includes images representing different TMA spots from the cancerous area of the prostate for each patient. ISUP scores were obtained during routine diagnostics, and tissue samples were sectioned at a thickness of 10µm, in line with the standard digitization protocol of the UKE.

### UKE-long

In the UKE-long sub-dataset, used for testing of the BASE and PCAI models, each image represents a different TMA spot from the cancerous area of the prostate for every patient. ISUP scores were determined during routine diagnostics. Tissue samples were stained with Hematoxylin and Eosin for an extended duration of 40 minutes and 10 minutes, respectively, nearly ten times the regular staining time. This experimental sub-dataset contains 1,667 images.

The datasets are split stratified by event indicator and, since patients can have multiple TMA spot images, we strictly separate patients across data splits. This means that TMAs of the same patient are present in either the training or test data but never in both. An overview of the patient characteristics for the sub-datasets can be found in Table S1.

### Prostate Cancer Biorepository Network TMAs

Two additional TMA datasets from the Prostate Cancer Biorepository Network (PCBN) ^18^ in the USA, collected at the New York Langone Medical Centre (NYU) and the Johns Hopkins Hospital in Baltimore (JHU), are included as depicted in Fig. 1B and discussed in detail in the following. Note that every patient received RP treatment for all PCBN datasets, similar to the UKE-high-variance TMA dataset. As for the UKE-high-variance TMA data, expert pathologists ISUP graded the whole prostate and this annotation was used for the individual TMA spot.

#### NYU

The TMA cohort from New York University (NYU), used for testing of the BASE and PCAI models, contains a total of 204 unique patients arranged in four TMA blocks. ISUP grading is assessed on a patient level and no additional grading details like the number of pathologists are provided. This dataset includes four TMA blocks of tissue spots (0.6 mm in diameter) from prostatectomy specimens. These spots were sectioned at 5µm in contrast to 2.5µm in the internal UKEhv dataset (with the notable exception of UKE-thin and -thick). The TMA block images were digitized using an Aperio scanner with a magnification of 20x (0.5µm per pixel) and cut into individual images of size 1817x1817 pixels using QuPath ^19^. Spots showing non-neoplastic tissue were excluded. After preprocessing and filtering, this work integrated 515 images of 161 patients with a median of 3 images per patient.

#### JHU

The TMA prostatectomy samples from the Johns Hopkins University (JHU), used for testing of the BASE and PCAI models, were derived from two datasets named “Case Natural History of Prostate Cancer” (6 TMA blocks) with 235 patients and “Case PSA Progression” (16 TMA blocks) with 726 patients. All patients underwent radical prostatectomy. ISUP grading is assessed on a patient level and no additional grading details like the number of pathologists are provided. In contrast to the other TMA datasets, the endpoint definition of this dataset in terms of event duration is only accessible in a granularity of years instead of days. These two datasets also contain rich metadata information like age, body mass index, race, local recurrence, etc. that was disregarded in this work’s analysis. Moreover, we extended the aforementioned event indications by salvage treatment, leading to a censoring rate for this dataset of under 1%. This means that this cohort can be considered to be biased towards unhealthy individuals. Also, it expresses the highest ratio of M1 (37.2%) as well as N1 (18.6%) patients, which is expected since the “Case PSA Progression” patients all had BCRs. This is further emphasized by the highest relapse rate of JHU patients among all TMA spot datasets in the overall KM curves (data not shown). The TMAs were sectioned with a thickness of 4 *µ*m and scanned with a Ventana DP2005 and a Hamamatsu NanoZoomer XP6 scanner. For integration, the 22 TMA block images were cut into individual spot images of size 3200x3200 pixels at a magnification of 40x (0.25 µm per pixel) using Qupath. After preprocessing, this work integrated 3,575 TMA spot images that show prostatic adenocarcinoma from 879 patients, with a median of four images per patient.

### UKE-sealed TMAs

The UKE-sealed TMA dataset, used for testing of the BASE and PCAI models, contains 826 patients and 4,095 images with a maximum of 10 images per patient. This dataset is special, in that it contains spot-level quantitative Gleason grading that included the percentage of Gleason 3, 4, and 5 patterns from GS, as opposed to the prostate-level annotations for spots of all other TMA datasets in this study. The information of quantitative Gleason grades was subsequently used to calculate the spot-wise IQ-Gleason, the currently best-performing clinical PCa grading system, aggregated as the mean or maximum over all images of a single patient^15^. UKE-sealed is therefore the only TMA dataset where we can objectively compare the predictive performance of our algorithm to the ISUP grading, since both utilize the exact same images and information. The name UKE-sealed stems from the fact that the access to all patient, metadata, and outcome information was and is restricted exclusively to the department of Pathology of the UKE. Also the evaluation of TMA spot predictions were conducted exclusively by the department of Pathology of the UKE. Hence, this dataset provides an objective point of performance comparison for the BASE and PCAI models.

### Malmö (MMX) biopsies

The MMX biopsy dataset from Malmö, Sweden, was used for the testing of the BASE and PCAI algorithms and contains 716 patients originally collected by Saemundsson et al. ^20^. Patient-level ISUP scores were obtained during routine diagnostics. To further allow for an image-level comparison against the BASE and PCAI models, three individual pathologists assigned image-level ISUP grades to all slides independently and blinded from any additional patient information. The biopsy-level ISUP grades provided by the three pathologists of two centers (Aachen and Uppsala) showed an interrater agreement Fleiss kappa of 0.199. The authors removed all patients with no or less than 2 mm of total cancer in their biopsy, missing follow-up information, inadequate RNA quality, and those that had already developed metastases at the time of diagnosis. Furthermore, for usage in this work, patients that were censored within the first five years as well as images with insufficient quality were removed. In total, 269 patients with 578 images were included in this work, with up to eight images for a single patient. The median survival and follow-up time for the remaining patients is 38 and 106 months respectively. Notably, the patients’ mean age in this dataset is 4-8 years older than the other datasets. Also, those patients show the highest average PSA values as well as a high variance with 19.9+-44.5 ng/mL. The time-to-event measurement begins with the biopsy date, leading to longer observed time spans in comparison to the TMA datasets, where the reference point is the date of RP. The images of this dataset were digitized using Hamamatsu and Ventana scanners at 40x magnification resulting in individual slide images with a resolution of 0.23µm per pixel. Image widths and heights vary but consist of up to hundreds of thousands of pixels for the long side of a biopsy.

### Uppsala (UPP) biopsies

The UPP biopsy dataset from Uppsala, Sweden contains 2,611 unfiltered images of 440 patients from the SPROB20 image dataset that was enriched by patient endpoint information and was used for the testing of the BASE and PCAI algorithms ^21^. Since some patients in this dataset have had multiple biopsies taken, this work only considers biopsy images from the latest patient visit and excludes all earlier biopsies. ISUP scores were obtained from the pathology report of the fusion biopsies during routine diagnostics. Additionally, patients without an assigned ISUP grade, as well as patients with incomplete or conflicting treatment and follow-up information were excluded from this dataset. In total, 683 images of 123 patients of this dataset are included in the evaluation of PCAI, with up to 10 images per patient at point of biopsy. The UPP biopsy slides were sectioned at a thickness of 4-5 µm and digital images were obtained from a Hamamatsu scanner on a magnification of 40x (0.25 µm per pixel). Since this cohort contains patients from a pilot study for MRI-guided acquisition of prostate biopsies the number of missed biopsies may be different, higher or lower, than it would have been if the conventional procedure had been used.

### PANDA data

The PANDA dataset contains biopsy slides and corresponding tissue and cancer annotations of pathologists. This dataset was only used to train the CI model to find cancerous areas of biopsy images and not for endpoint prediction. PANDA is one of the largest publicly available whole slide image (WSI) datasets in the world with 10,616 provided biopsy slides with slide-level primary and secondary Gleason score as well as ISUP annotation from 2,113 patients. It was published in the Prostate Cancer Grade Assessment challenge and a part of the International Conference on Medical Image Computing and Computer Assisted Intervention (MICCAI) in 2020 ^22^. The training and validation data for this challenge is provided by two centers, the Karolinska Institute in Stockholm, Sweden (5,456 WSIs) and the Radboud University Medical Center in Nijmengen, Netherlands (5,160 WSIs). The PANDA dataset is used to train the cancer indicator based on 9,554 training- and 1062 test WSIs with 3.94 and 0.51 million extracted patches respectively (see Fig. S1), which utilizes the expert annotations to classify extracted tissue patches into cancer containing or benign tissue. The main purpose of the cancer indicator is to function as a patch selector for the PCAI model to identify the most relevant patches for risk assessment from up to tens of thousands of patches per biopsy. In addition, the CI increases the interpretability of PCAI’s results by highlighting decision-relevant cancerous areas.

### PCAI design rationale

PCAI is an end-to-end risk assessment model for histopathological prostate cancer data, aimed at actual implementation in a real-world environment (Fig. 3). PCAI is built upon four pillars of clinical applicability, which formed the basis of our design decisions.

**Figure 3.**
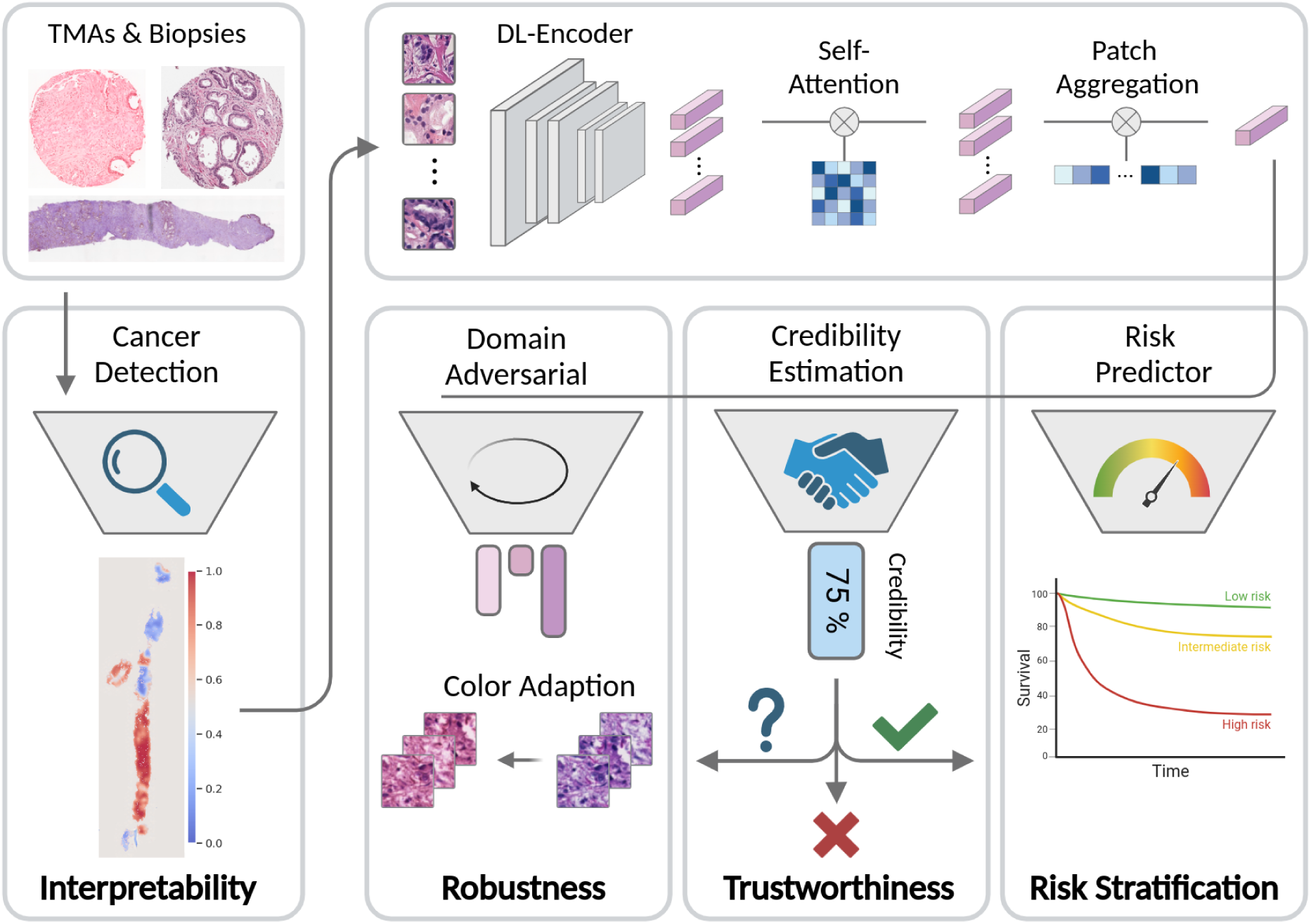
Overview of the PCAI PCa cancer grading algorithm. PCAI is based on four pillars of clinical applicability: It can process arbitrarily sized TMA-spot and biopsy images. A separate cancer indication network highlights highly cancerous regions on the input images, providing visual interpretability. The most relevant patches are then processed in the deep-learning network and used for cancer grading that exceeds stratification based on expert-assigned ISUP. A credibility estimation setup outputs a credibility score with every risk prediction that adds a quantifiable measure of trustworthiness. Finally, a domain adversarial training regime as well as a credibility-guided color adaptation setup contribute to the model’s robustness across five unseen TMA-spot and biopsy datasets.

First, the model’s risk prediction performance should exceed that of the subjective five-tier ISUP scoring system. We hypothesized that this is best possible if the model learns how to predict objective patient outcomes over time instead of replicating a subjective ISUP grade by “experts”. To this end all datasets used in this work contain at least 5 years of follow up information for all patients. PCAI then grades the cancer by predicting a potential disease relapse in the future.

A leading cause for model prediction errors are variations in the processing of histopathological samples. To render PCAI robust to these changes, we hypothesized that domain adversarial training ^23,24^ on our large and heterogeneous UKEhv dataset will result in stable predictions across unseen datasets and domains that reflect the variance encountered in everyday clinical practice (details in Supplemental Information).

Even though PCAI has been trained and optimized for stable predictions across different sample processing protocols, it might still encounter histopathological slides of e.g. bad quality, for which it cannot provide a reliable grading. A relevant feature for PCAI is therefore the notion of confidence or trustworthiness via credibility estimation, not unlike a human expert that is uncertain about the grading of a particular sample and asks for a second opinion (details in Supplemental Information).

With the aim to stabilize PCAI on images where it shows a low confidence, we further introduce a credibility-guided color adaptation procedure that maps the color scheme of low-credible samples to that of the model’s training distribution. As we later show, this additionally improves overall performance and robustness. If probes are still problematic after color adaptation, the grading of them can be deferred to the pathologist (Fig. S4A-B).

A very relevant feature of PCAI is its interpretability, which lets human experts understand and trust model predictions. PCAI achieves this via its cancer indicator module, which highlights and selects cancer regions of the sample with an accuracy of over 95% as assessed on the PANDAs dataset.

With these design features in mind, we first derive a baseline (BASE) model, trained only on the single internal data domain UKE-first, containing the most predictive TMA spot per patient, according to the collecting pathologist. We then utilized UKE-first, UKE-second, and UKE-scanner datasets of the UKEhv cohort and applied several techniques to increase robustness (domain adversarial training, color adaptation), trustworthiness (credibility estimation), and interpretability (cancer indication, risk groups). The combination of these techniques represents our final risk prediction model PCAI (Fig. 3, S1B-C). PCAI leverages the morphological features learned by training a CNN-based neural network architecture (EfficientNet ^25^) with postoperative TMA-spot image data (UKE-first, UKE-second, and UKE-scanner) and corresponding patient follow-up information to allow for a valid risk prediction on clinically more relevant, preoperative biopsy images. As is common practice when applying deep learning models on gigapixel histopathological images, we automatically segment the relevant tissue regions as described below of every image and crop it into equally sized patches of 128x128 pixels at 20x magnification. The relevant patches for each image are then fed into the network and transformed into a lower dimensional representation by the encoder module (Fig. 3, S1). These are further cross-correlated in the self-attention ^26^ module, to account for dependencies across the image and to create context-aware embeddings from every patch. The subsequent attention-based aggregation module ^27^ assigns a score of relative importance to every patch and combines them into a single representation. Finally, the fully-connected risk prediction head infers the patient’s probability of having a relapse in the first five years after sample acquisition from the combined patch embedding. The predicted probability represents our PCAI risk score. Notably, utilizing the large number of smaller TMA spot images allowed us to train our network in an end-to-end fashion and adapt all parts of our pipeline specifically to the task at hand, without the need to rely on pre-trained models or self-supervised methods, as is often the case in histopathological deep learning applications. During inference, PCAI can process arbitrarily sized images due to the patch-based image processing and aggregation in our model.

Details of the full algorithm, including deep-learning architecture, hyperparameter and training details are provided in the Supplemental Information.

## Results

### The effect of data variation on the BASE model

Data variation can significantly reduce the performance of AI-based decision systems, while it seems to have minor effects on human experts. This problem exists across biomedical domains, ranging from genomics to image processing and beyond ^17,28,29^. Data variation of histopathological slides stems in part from differences in the formalin fixation and paraffin embedding of tissue, sectioning, staining, and slide scanning.

In this work, we aimed to systematically investigate the effect of data variation on AI-based PCa grading performance, using the UKEhv dataset. The dataset contains TMAs processed with the standard protocol (UKE-first), using a different lot of reagents and core (UKE-second), an extended staining (UKE-long), thinner (1 µm, UKE-thin) and thicker (10 µm, UKE-thick) sectioning (2.5µm standard), and a different scanner (UKE-scanner) (Fig. 2, Table S2). Notably, the dataset contains all protocol variations for a subset of the same 1,537 patients, taken from the same RP sample, constituting an ideal basis for the evaluation of data variation on model performance. As can be observed in Fig. 4A, the UKE high-variance datasets show marked differences in the visual appearance, which is further confirmed in Fig. S3.

**Figure 4.**
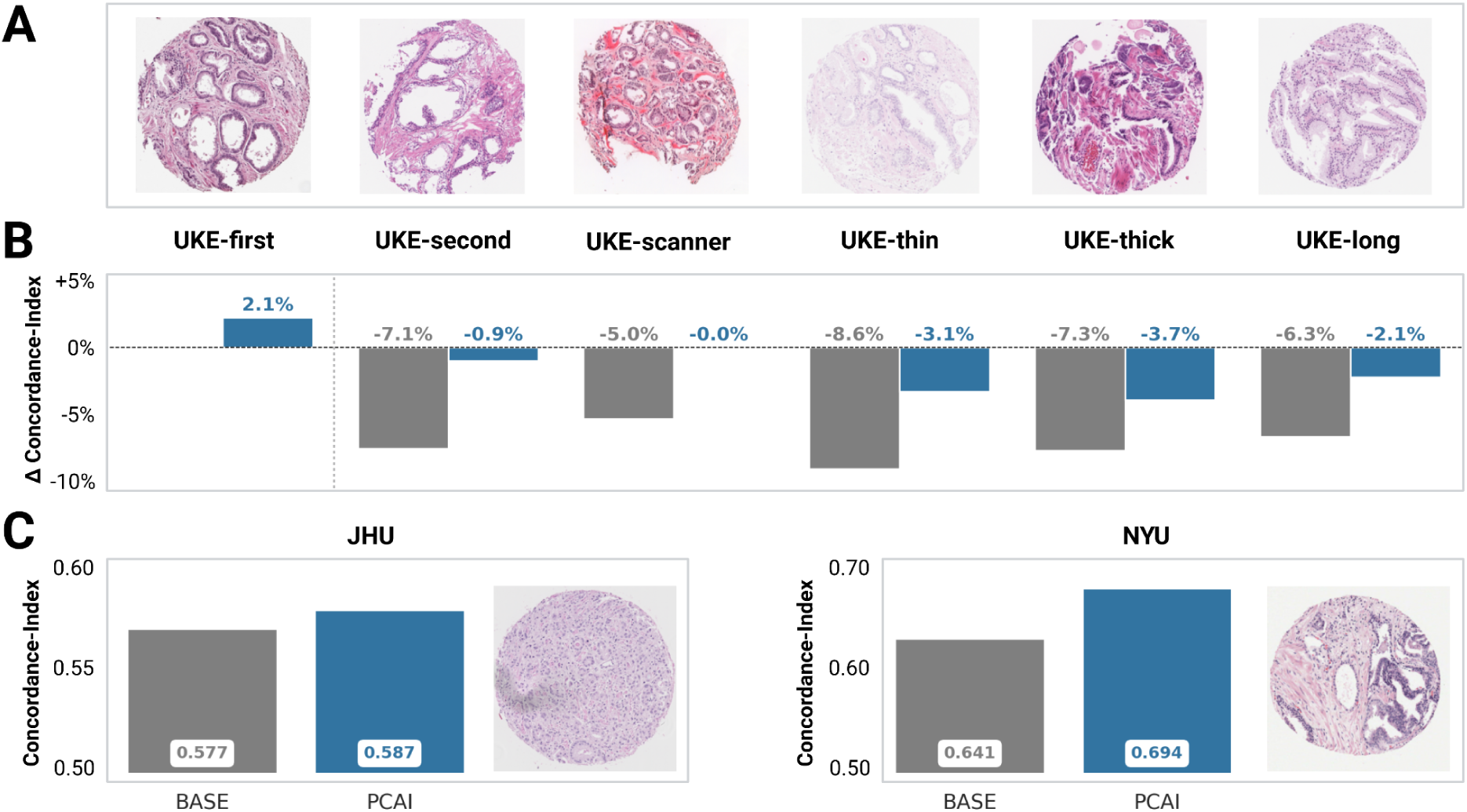
The effect of data variance on model performance. **A** Exemplary images for each of the UKEhv sub-datasets **B** Difference in C-Index of PCAI (blue) and BASE (gray) compared to that of BASE on UKE-first for the same overlapping patients (n=1537) with one image in each sub-dataset. **C** Absolute performance in terms of C-Index of PCAI and BASE on the JHU and NYU datasets.

First, we trained the BASE model (Fig. S2A) on the UKE-first training set and evaluated its performance on all six UKEhv test datasets. As is standard in all of our analyses, the training and test datasets did not contain any overlapping patients to avoid overestimation of model performance. Although the BASE model is trained on the very large UKE-first training dataset of 8,123 images and 8,123 patients, a significant decrease in prediction performance is seen across all data variations. While the BASE model achieves a C-Index of 0.645 on the UKE-first dataset, the performance drops significantly by 5 (UKE-scanner) to 8.6 (on UKE-thin) percentage points on the same patient population for other TMAs in the UKEhv dataset (Fig. 4B).

These results strongly indicate that AI-based models, even when trained on large datasets and using image augmentation ^30^, have significant difficulties with data variations they were not trained on. Standard operating procedures for histopathology vary with the location but also over time at the same medical center, which constitutes a central problem for the robustness and fidelity of AI-based PCa grading systems.

### Conferring algorithmic robustness to data variation

An algorithm can never be trained on all current and future data variations. It is therefore pivotal to stabilize AI-based PCa grading by using algorithmic modifications such as domain adversarial training (Fig. 3). Even when stabilized algorithmically and trained on large datasets, a model might still encounter probes that are difficult for it to grade. A relevant feature for a model is therefore the notion of prediction confidence or trustworthiness, not unlike a human expert that is uncertain about the grading of a particular sample and asks for a second opinion (details in Supplemental Information). A model that assesses the confidence and estimates the credibility of its predictions is able to fix problematic probes using color adaptation or defer them to a pathologist (Fig. S4A-B). Lastly, the addition of cancer maps allows for an interpretation of the prediction results by a medical expert.

In accordance with these deliberations we implemented algorithmic modifications for robustness, credibility, and interpretability into the BASE model, naming it PCAI (Fig. 3 & S2C), and again assessed the performance on the UKEhv dataset. We trained PCAI on the sub-datasets UKE-first, UKE-second, and UKE-scanner, consisting of 19,883 images and 6,937 patients, and evaluated the performance on the same test datasets as the BASE model.

We first assessed the impact of credibility-guided color adaptation on the PCAI model, which increased the average 5-year AUROC across all UKEhv datasets as well as the US American JHU and NYU TMA datasets by 0.9 percentage points on average, with up to 4.5 percentage points on the NYU dataset (Fig. S4C).

Furthermore, PCAI showed significantly increased grading performance across all data variations. In brief, the performance of PCAI on the UKEhv test datasets increased by 2.2 percentage points on UKE-first (0.645 to 0.667), 6.2 on UKE-second (0.574 to 0.636), 5.0 on UKE-scanner (0.595 to 0.645), 5.4 on UKE-thin (0.560 to 0.614), 3.5 on UKE-thick (0.573 to 0.608), and 4.2 on UKE-long (0.582 to 0.624) over BASE (Fig. 4B). We next evaluated the performance of the BASE and PCAI models on the NYU and JHU datasets. Again the performance of PCAI increased by 1 percentage point for NYU (0.577 to 0.587) and by 5.3 on JHU (0.641 to 0.694) over BASE (Fig. 4C).

These results strongly suggest that algorithmic modifications for robustness and credibility, in conjunction with a high quality training dataset, can make AI-based PCa grading algorithms robust to a broad spectrum of data variations.

### Human interpretation of PCAI’s predictions

It is pivotal that AI-based clinical decisions support systems are interpretable by human experts. Interpretability allows the expert to trust or ignore a model prediction. PCAI delivers at least two highly interpretable results. First, PCAI’s cancer indicator provides visual cues on images as to the location of cancerous areas (Fig. 5A-B). Second, PCAI can distinguish 7 different risk groups in order to transform the continuous cancer grade score into categories that may be more appreciated for clinical interpretation than continuous data (Fig. 5C-D).

**Figure 5.**
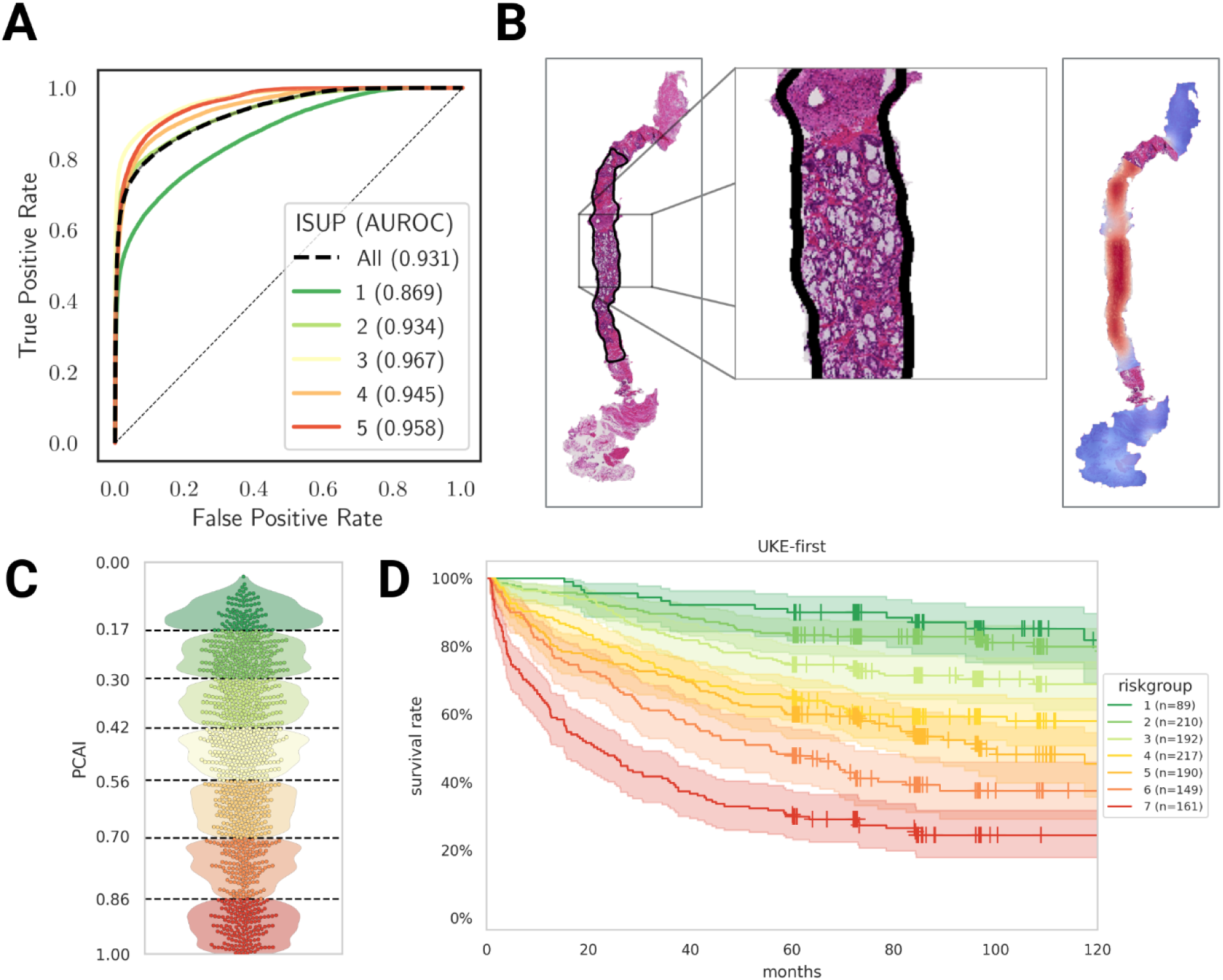
Interpretation of PCAI. **A** Overall and per ISUP patch-wise cancer classification ROC of the cancer indicator on the PANDA test data. **B** Human annotated outline (left) and cancer-indicator-generated heatmap (right) of an exemplary PANDA biopsy sample where blue shaded regions show healthy tissue and red shaded regions contain cancer. **C** PCAI prediction distribution and risk groups for the UKE-first test data. **D** KM curve for stratified predictions of UKE-first that shows clear separation among the risk groups.

In detail, the cancer indicator allows for patch-wise prediction of cancer probability. The cancer indicator was trained on the PANDA dataset and achieves an AUROC of 0.94 on the PANDA test data (Fig. 5A). Interestingly, it can be observed that ISUP group 1 achieves a slightly lower AUROC (0.869) as compared to all other ISUP groups (AUROC >0.930). The cancer indicator enables us to create cancer probability heatmaps on all images, which provide visual means of interpretability that can also be used as an automatic quantitative indicator of how much cancerous tissue is present in the images (Fig. 5B). Especially on the biopsy images, where we use the cancer indicator to focus PCAI on the relevant cancerous regions, this transparently highlights salient regions that contributed to the final PCAI risk score.

PCAI’s cancer grading is a continuous score from 0 (low risk) to 1 (high risk), which poses the challenge to define the appropriate clinical decisions for a given score. To potentially enhance the interpretability of the predicted risk score, we derive 7 statistically distinct risk groups from the UKEhv training data by using k-means clustering (Fig. S5). The Kaplan-Meier curves of the UKE-first test data show a clear separation of the patients in our proposed risk groups (Fig. 5D, S6).

In summary, we equipped our risk prediction model PCAI with the necessary tools to provide robust (domain adversarial training, color adaptation), trustworthy (credibility estimation) and interpretable (cancer indication, risk groups) predictions, laying the foundation for actual clinical application. We now set out to evaluate if PCAI can rival or exceed the current clinical standard of ISUP ratings.

### PCAI surpasses expert ISUP grading on TMAs

A main aim of the patient outcome-based PCa grading prediction of the PCAI model lies in its potential ability to exceed the current five tier ISUP grading. We therefore assessed the performance of the BASE and PCAI models on the UKE-sealed dataset, which contains spot-level ISUP grading from UKE pathologists. In addition, the UKE-sealed TMA spots were graded using the IQ Gleason (GIQ) grading system, although 0,6mm TMA spots are not very well suited to assess percentages of Gleason patterns. The GIQ is currently one of the best performing grading systems for PCa histopathology ^31^. In the case of multiple TMAs for a single patient in the UKE-sealed dataset the average GIQ was calculated as the patient-level prediction. Similarly, the BASE and PCAI model were evaluated taking the mean aggregated image-wise predictions and compared to the worse ISUP grade. On UKE-sealed, PCAI exceeds BASE performance by 3.2 percentage points (C-Index of 0.712 to 0.744, Fig. 6A). Moreover, PCAI outperformed the five-tier ISUP grading by 3.1 percentage points in C-Index (Fig. 6A). Comparably, PCAI exceeds BASE performance by 3.4 percentage points regarding AUROC (0.780 to 0.746, Fig. 6B). PCAI outperformed ISUP grading by 2.3 percentage points in AUROC (Fig. 6B). Importantly, PCAI performed comparably to the GIQ grading (0.1 percentage points better in C-Index and 0.7 percentage points worse in AUROC). In this context it is interesting to note that the BASE model always performed worse than ISUP and GIQ grading. These results indicate that the algorithmic adaptations in PCAI resulted in above ISUP grading in C-Index and AUROC on TMAs.

**Figure 6.**
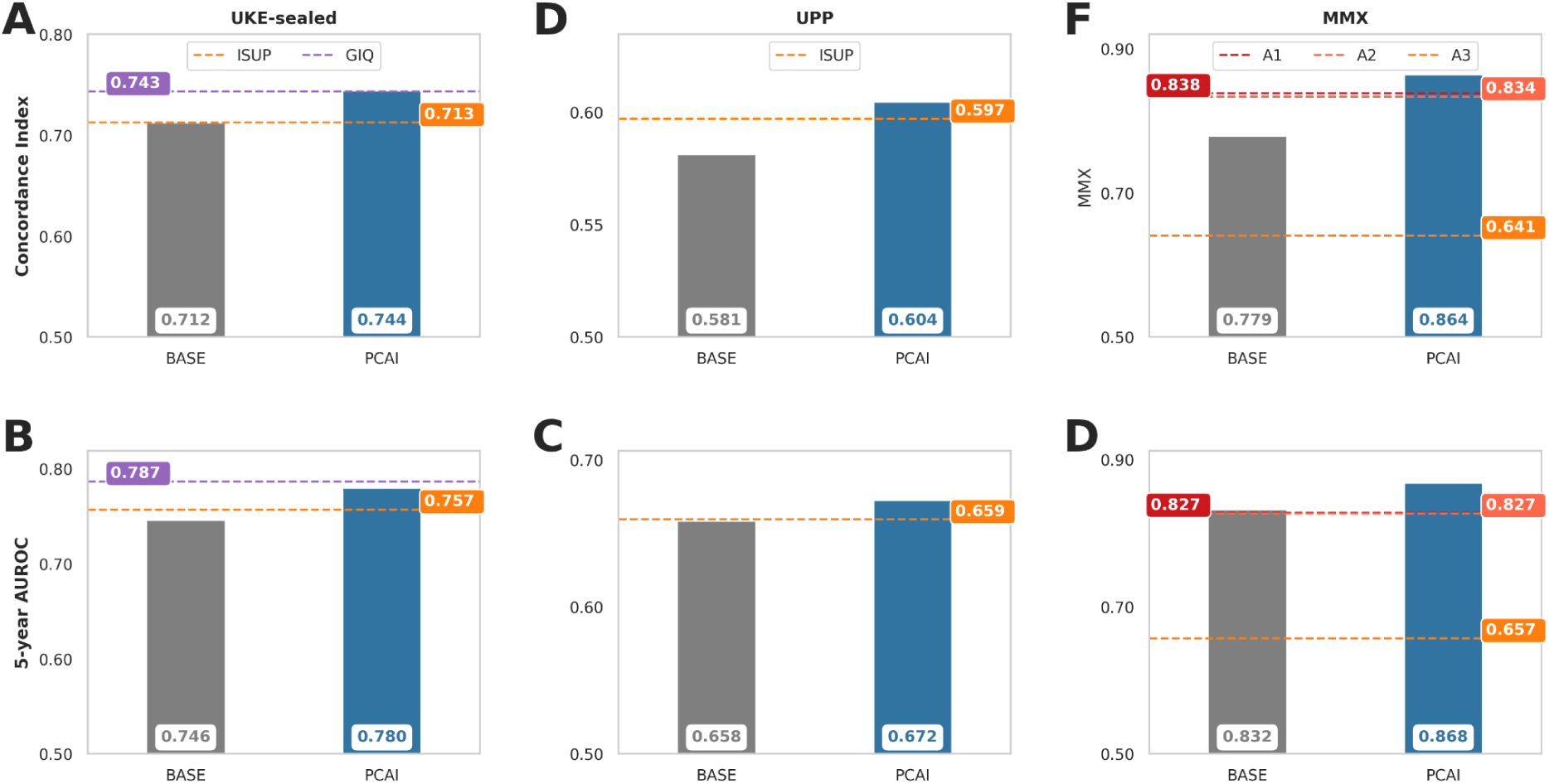
PCa grading performance of human experts and the BASE and PCAI models. Performance of BASE (gray) and PCAI (blue) in terms of C-Index and 5-year relapse AUROC on the UKE-sealed TMA spot dataset (**A**, **B**) and the UPP (**C**, **D**) and MMX (**E**, **F**) biopsy datasets. Performance of image-wise ISUP grade (annotated as ISUP, A1, A2, A3 in orange and red) or GIQ (purple) annotations are depicted by the horizontal dotted lines. To aggregate image-level predictions to patient-level, mean aggregation is performed on UKE-sealed while max aggregation is performed on the biopsy datasets (UPP and MMX).

### PCAI surpasses expert ISUP grading on biopsies

The litmus test for PCAI is whether its robustness and performance extends to clinical, preoperative biopsy PCa samples, which are imaged as significantly larger whole slide images (WSI) than TMA spots. For this, we evaluate PCAI on the two biopsy cohorts from two clinical centers in Sweden, UPP (123 patients and 683 images) and MMX (269 patients and 578 images). To focus PCAIs risk prediction on relevant tissue areas, we preselect the 100 most likely cancerous patches per whole slide image using PCAI’s CI module. If multiple WSIs per patient were available for a single biopsy, the maximum risk score across images was used as the patient-level prediction.

PCAI achieved a C-Index of 0.604 on the UPP dataset, which is 0.7 percentage points higher than ISUP (0.597) and 2.3 percentage points higher than BASE (0.581) (Fig. 6C). Similarly, PCAI reached an AUROC of 0.672 on the UPP dataset, which exceeds ISUP (0.659) by 1.3 and BASE (0.658) by 1.4 percentage points (Fig. 6D).

On the MMX data, PCAI achieved a C-Index of 0.864, 4.7 percentage points higher than ISUP (0.817) and 8.5 percentage points higher than BASE (0.779). Using AUROC as a measure of performance, PCAI (0.868) exceeded ISUP (0.813) predictions by 5.5 and BASE (0.832) predictions by 3.6 percentage points.

To obtain a notion of human inter-rater variability and assess significance, we also compared model performance to the image-wise ISUP grading of three highly skilled pathologists from Germany and Sweden. In C-Index, PCAI (0.864) significantly exceeded the performance of expert ISUP grading (A1: 0.838, A2: 0.834, A3: 0.641, mean: 0.771) by 9.3 (mean) percentage points (Fig. 6E). This holds also true for AUROC, where PCAI (0.868) significantly surpasses expert ISUP (A1: 0.827, A2: 0.827, A3: 0.657, mean: 0.770) grading by 9.8 (mean) percentage points (Fig. 6F). Notably, PCAI scores consistently high for every relapse cutoff point (Fig. S7).

Taken together, these results on biopsy-derived whole slide images substantiate our findings on UKE-sealed TMAs, highlighting the importance of our algorithmic adaptations to exceed the five-tier ISUP-level cancer grading on multiple, highly variable datasets.

## DISCUSSION

In this work we have developed PCAI, a robust, trustworthy, and interpretable PCa risk assessment model with state-of-the-art performance, trained patient outcome on one of the largest TMA-spot datasets used in the literature to date. A highlight of PCAI is surely that it uses patient outcome as a cancer aggressiveness proxy, a step towards ‘more objective’ cancer grading away from human made grading systems. PCAI generalizes on unseen TMA spot data of three different centers. On the UKE-sealed TMA test dataset, PCAI outperforms the five-tier ISUP grading and yields comparable results to the GIQ score. In addition, PCAI outperforms ISUP grading on two PCa biopsy cohorts from two clinical centers in Sweden. On the UPP dataset, PCAI outperforms the ISUP grade in terms of C-index and AUROC. To further demonstrate the predictive performance of PCAI over ISUP grading, we incorporated image-wise ISUP annotations of three (experienced) pathologists from different clinics on the MMX dataset and evaluated our risk score against those. PCAI outperforms ISUP grading significantly on the mean prediction performance in C-index and AUROC and exceeds ISUP predictions irrespective of the individual pathologists in every individual comparison, reaching an AUROC of 0.87. The obvious superiority of PCAI over pathologists is driven by the inherent limitation of a five-tier prostate cancer grading system such as the current ISUP approach. Several large-scale studies from our group have demonstrated that the traditional Glaseon grading can be markedly improved by taking into account the percentages of Gleason 4 and Gleason 5 patterns in a tumor ^2,31^. That the PCAI achieved a comparable performance to the IQ Gleason on 4095 TMA samples may argue that the performance of our PCAI comes close to the best possible use of Gleason pattern evaluation. As digital image analysis is not limited to the gland architecture but can also take into account other potentially prognostic parameters such as nuclear features and stroma composition, further improvements of AI based prognosis assessment models should be possible.

Our adaptations to the BASE model to introduce more robustness, namely joint domain adversarial training, credibility estimation and color adaptation, showed a consistent improvement in predictive performance across all datasets. Especially on the JHU, UPP and MMX cohort, these adaptations provided the decisive edge to surpass the prediction value of ISUP grading. The high separability of the PCAI risk groups derived from the internal UKE test set, which is also visible in the respective Kaplan-Meier curves, proves a strong correlation of PCAI’s output score with the actual patient endpoints. Interpretability is further enhanced by the reported credibility score for every sample, which provides a notion of trustworthiness of our model’s prediction.

The recent progress in AI-based Gleason grading and PCa aggressiveness prediction is considerable, resulting in several cancer detection and grading systems that have been approved for clinical use. Most systems, however, have been trained on human annotations for cancerous regions and cancer grade according to the Gleason system. This poses two limitations to the algorithms, as they are bound by subjective expert decisions and the quality of the grading system they mimic. In this work we provide evidence that an AI-based model can consistently outperform expert-based five-tier ISUP grading in C-index and AUROC when trained on objective patient outcome. PCAI’s quantitative cancer aggressiveness score can be clustered into grades that include more than 5 risk groups, to allow for easier interpretation in a clinical setting.

In this context it is to be noted that the chosen endpoints signify different disease progression states, as biochemical recurrence should precede a metastasis and cancer-related death. The algorithm predicts an adverse course of the disease based on the first available adverse endpoint that was documented in the respective dataset. In the vast majority of patients the event was based on the occurrence of a biochemical relapse (BCR). In patients where the time of BCR was not available, other endpoints that correlate with cancer aggressiveness were used for prediction, for instance metastasis, which accounted for only 69 of 8822 cases in the UKE dataset. While in this work we chose to combine these endpoints to include as many patients as possible, future versions of PCAI with stratified endpoints might show further performance improvements.

Before decision support systems such as PCAI can be generally adapted in clinical routine, they have to prove their ability to handle the large data variability and give an accurate measurement of their confidence in the results. Although the first support systems have been approved by regulatory agencies, their application is still limited by the data they were trained on. Our work is unique, because we systematically evaluated the robustness of AI-based histopathological PCa prediction when faced with variations in data acquisition and processing. Even when trained on thousands of patients and tens of thousands of images, models show significantly diminished predictive performance when subjected to variations in slide thickness and staining time. The introduction of domain adversarial training and credibility-guided color adaptation, in conjunction with training data of higher variability, resulted in robust model performance in C-index and AUROC across multiple high-variability and three external test datasets. It is important to mention that PCAI was exclusively trained on TMA data and generalized with state-of-the-art performance to the PCa grading of external test biopsies. These results, however, do not imply that PCAI cannot fail on yet unseen data, which is why credibility estimation by conformal prediction is so important. By utilizing a measure of trustworthiness, PCAI is able to confer problematic biopsies to the pathologist. It is perplexing to realize the ease with which pathologists seem to deal with data variation and how detrimental it can be for best-of-breed AI models. While our results present solutions on how robustness to data variation and credibility estimation might be achieved, they also highlight a lingering weakness of AI-based systems, in general. The negative impact of data variation on AI-based clinical decision support systems requires further attention and future solutions should include collaborative data sharing strategies, the establishment of robust data standards, the development of further algorithmic strategies, and the leveraging of larger and more heterogeneous training datasets.

In conclusion, this work highlighted some salient problems with AI-based grading of PCa biopsies and offered some solutions to further increase the quality and robustness of existing algorithms.

### Computational hardware and software

The project was implemented using python 3.10 with pytorch-lightning 1.8.2. The PCAI, baseline and CI models were trained on a NVIDIA Quadro RTX 8000 with 48GB GPU memory and an Intel(R) Xeon(R) Silver 4214 CPU. A distilled version of the code is available at https://github.com/imsb-uke/pcai/.

## Data availability

Data from Prostate Cancer Biorepository Network (PCBN), namely the JHU and NYU datasets in this work are available upon request at https://prostatebiorepository.org/.

The PANDA dataset with corresponding GT segmentation masks is available on the challenge website at https://panda.grand-challenge.org/data/.

The UPP dataset images are publicly available under the name SPROB20 at https://datahub.aida.scilifelab.se/10.23698/aida/sprob20. However, the public version is anonymized and does not provide metadata such as endpoint information for the individual biopsies and patients.

The UKEhv, UKE-sealed, and MMX datasets are not publicly available.

## Author contributions

SB and GS initialized this work. SB, MZ, GS, EB, and PW conceptualized the project, algorithm, and computational analyses. SL, AD, PS, VGP, MH, MB, JB, PB, PW, AD, CB, MG, EB, GS, MZ, and SB supervised the work. GS, CB, PR, RB, and SvS helped grading some of the cohorts. GS and ML collected and aggregated the UKE cohort. PR, SL, MH, and AD collected and annotated the UPP cohort. FW, PF, ED, RK, NK, PW, AW, and MZ were responsible for implementing and running the algorithms and computational analyses. SB, FW, PF, MB, and MZ wrote the manuscript. PB, GS, and all other authors critically read and amended the manuscript.

## Acknowledgements

We would like to thank the IT of the Institute for Medical Systems Biology and the bAIome Center for Biomedical AI, Sven Heins, Sergio Oller, and Vadim Ustinov.

This work is supported by the Department of Defense Prostate Cancer Research Program, DOD Award No W81XWH-18-2-0013, W81XWH-18-2-0015, W81XWH-18-2-0016, W81XWH-18-2-0017, W81XWH-18-2-0018 and W81XWH-18-2-0019 PCRP Prostate Cancer Biorepository Network (PCBN)

JHU 726: We acknowledge the contributions of Dr. Angelo M. De Marzo and Helen L. Fedor from the Brady Urological Research Institute Prostate Specimen Repository at the Johns Hopkins University School of Medicine and Dr. Elizabeth A. Platz from the Department of Epidemiology at the Johns Hopkins Bloomberg School of Public Health for the generation of the ‘Case PSA Progression’ TMAs for the prostate cancer recurrence nested case-control study, funded in part by the Prostate SPORE Pathology Core (P50 CA58236), Oncology Tissue and Imaging Services at the Sidney Kimmel Comprehensive Cancer Center (P30 CA006973), and a DOD grant (DAMD 17-03-0273).

JHU 235: We acknowledge Drs. Patrick C. Walsh, Alan Partin and Misop Han of The Johns Hopkins University School of Medicine Brady Urological Research Institute for dataset curation for the 235 patient ’ Natural History of Prostate Cancer’ TMA.

PB is supported by the German Research Foundation (DFG, Project IDs 322900939, 432698239 & 445703531), European Research Council (ERC Consolidator Grant No 101001791), and the Federal Ministry of Education and Research (BMBF, STOP-FSGS-01GM2202C).

RK was supported by DFG FOR5068 P9 and the 3R initiative of the UKE, PF and MB by DFG SFB1286 SP02, and SB by EU E-rare MAXOMOD, the M3I excellence initiative of the UKE, and DFG SFB1192 B8 and C3. NK was supported by DFG SFB1192 B8 and AW, MZ, and JB by CDL FLIGHT of the University of Hamburg. ED was supported by DFG KFO306 and FW by DFG KFO296.

