## Supplements for "Robust, credible, and interpretable AI-based histopathological prostate cancer grading"

### Robust, credible, and interpretable AI-based Prostate cancer digital pathology with above expert detection and grading performance

#### Supplemental Information

##### PCAI model

The PCAI model describes our overall risk prediction algorithm. It combines the baseline neural network (**BASE**) with multiple adaptations for robustness, trustworthiness and interpretability, namely domain adversarial training (**DA**) on multiple internal data domains, the feedback loop of credibility estimation (**CE**) and color adaptation (**CA**) during inference, as well as the cancer indicator (**CI**) guided patch-selection for biopsies.

##### Preprocessing

Since histopathological images come in arbitrary shapes and sizes and contain a lot of redundant background pixels, we use a masking procedure to define the relevant tissue area in every image for usage in our network. In detail, we first create a tissue mask by separating foreground and background pixels using Otsu thresholding. In the second step, we create an anomaly mask by highlighting all foreground pixels with values outside a predefined deviation of the median of pixel values of the tissue area. This removes pen marks, blood or other undesired areas of the images, which are especially prevalent on the large biopsy images. A patch-based approach was used for our risk prediction network, as is common practice in digital pathology. For this, the images are further cut into equally sized patches of 128x128 pixels at 20x magnification based on the relevant tissue area defined by the masks. We refer to the entirety of *n* patches of an individual WSI as "patch bag". We then assign a binary label to each patch bag, indicating whether the patient experienced a relapse (defined as biochemical recurrence, metastasis or PCa-related death) in the first 5 years after examination.

**Data Splitting**

The three larger sub-datasets of the UKEhv data, UKE-first, UKE-second and UKE-scanner, are split into training, validation and test set (70/15/15), and the three smaller sub-dataset UKE-thin, UKEthick and UKE-long are split into validation and test set (50/50). The data is split stratified by the binary 5-year survival indicator. Patients that contribute images to multiple sub-datasets are strictly separated across data splits to avoid leakage. Final numbers per split slightly deviate from the initial percentages since some images were excluded after assigning the split due to the image filter criteria. This work uses the training set of the UKE-first data to train the BASE model and the training sets of the UKE-first, UKE-second and UKE-scanner data to train the DA model.

The remaining datasets UKE-sealed, JHU, NYU, UPP and MMX are only used for testing.

**BASE**

The baseline risk prediction network BASE is a binary classifier that assigns the probability of having a relapse in the first 5 years after examination to a bag of patches per image (Fig. S2A). Since the relapse information corresponds to the full patch bag and no ground truth for individual patches is available, information of patches inside one bag needs to be aggregated. This is referred to as multiple instance learning. In detail, we use the encoder part of EfficientNet-b0 to extract latent information of all n patches in a bag independently [25]. Next, a self-attention layer (SA), as proposed by Rymarczyk et al. [26], accounts for cross-dependencies between all patches of a bag. For every patch *i*, *n* attention weights are computed, resulting in the attention matrix $A_{SA} \in R^{n \times n}$. $A_{SA}$ contains information about the relevance of each patch i in relation to every other patch j and is multiplied with the incoming bag feature vector after the encoder. This creates context-aware embeddings from every patch. This bag of patch embeddings is further aggregated into a single latent representation in the attention-based multiple instance learning layer (MIL), as proposed by Ilse et al. [27]. For every patch $i$, one attention weight is computed, resulting in the attention vector $A \in R^n$. A softmax function ensures all weights sum to one. Multiplying $A$ with the incoming patch bag yields the aggregated representation of shape $1 \times L$. This method can be seen as a learnable weighted averaging function. Finally, the risk classification head, consisting of two fully connected layers (1280 → 100 → 2 neurons) predicts the probability for both classes, using softmax activation function. The predicted probability for class 1, corresponding to having a relapse prior to five years, represents our final risk score. Fig. S2A depicts a schematic of the BASE architecture.

The BASE model is derived exclusively from the UKE-first dataset. We train our network end-to-end using 100 randomly over- or undersampled patches per image with a batch size of 16, Adam optimizer and a learning rate of 2.75e-06 for a maximum of 200 epochs, with early stopping on the 5-year AUROC of the UKE-first validation split data. Dropout rate and stochastic depth of the EfficientNet backbone are both set to 0.34. The static number of 100 patches allowed for training with batch sizes > 1 and was chosen to be close to the median number of valid tissue patches across samples in the dataset. Patches were further randomly transformed with AugMix augmentation before input to the network to increase data variance and robustness [30]. We use class-weighted cross-entropy as our loss function. Hyperparameters were optimized for maximum 5-year AUROC on the UKE-first validation split data using a Bayesian search paradigm. During inference, all valid patches per image and no AugMix augmentations are used. If multiple images of any type are available for a single patient and examination, we aggregate by taking only the highest risk score predicted by our model as the final patient score.

**Domain adversarial training**

For the DA training, the BASE architecture is extended by a domain discriminator head, as well as a gradient reversal layer (GRL) between MIL layer and domain discriminator (Fig. S2B) [23]. We extend our UKE-first training dataset by data from the UKE-second and UKE-scanner sub-datasets and assign a secondary domain label to every image, indicating from which sub-dataset it originates. We then train our DA model in a dual-task manner, where the domain discrimination head aims to correctly predict the sub-dataset a given image stems from. The key concept of domain adversarial training is then applied through the GRL, which serves identity function during the forward pass, however flips the sign of the gradient during backpropagation. This enforces adaptation of the weights of the shared network part, consisting of encoder, SA and MIL layer in the exact opposite direction of the domain discrimination loss. This leads to the desired adversarial game between an consistently improving domain discriminator head and the shared network part, which provides latent representations of the data that contain increasingly less domain-specific information. Since the main task of binary 5-year relapse classification is trained in parallel, this allows the network to provide accurate risk predictions on domain invariant features. This method is inspired by Wilm et al., who prove the positive influence of DA for mitotic figure detection on histopathological images [24].

We optimize an additional parameter $\lambda$ that controls the influence of our domain adversarial loss, resulting in the overall loss function as a sum of the cross-entropy loss of the risk predictor and the cross-entropy loss of the domain discriminator as

$$L_{total} = L_{risk}^{CE} + \lambda \cdot L_{domain}^{CE}.$$

Training procedure in the DA model is analogous to the baseline model, though here a learning rate of 9.87e-07, dropout rate of 0.5 and stochastic depth of 0.5 is used. Data from the UKE-first domain was fed twice per epoch to put a stronger emphasis on the data containing the most representative spot per patient. We further perform early stopping as well as hyperparameter optimization on the combined 5-year AUROC of the validation splits of all UKEhv subdomains.

**Credibility estimation**

To be applicable in an actual clinical setting, the predicted risk score should be accompanied with a notion of trustworthiness that quantifies how certain the model is when predicting on a given image (Fig. S2C). For this we introduce the concept of credibility by computing a score for every unseen sample based on the distance to the learned distribution of the model. The underlying assumption is that samples that differ strongly from the data seen during training should receive a lower credibility score than those close to the learned distribution, independent of the actual predicted risk score.

In detail, we measure the Mahalanobis distance $d_M$ between the latent representation of an unseen sample in the output of the MIL layer to the center of the latent representation of all training samples. To further transform the Mahalanobis distance $d_M$ to the training center into a normalized representation of model uncertainty, ideas from the concept of conformal prediction (CP) are employed [32]. CP is a post-hoc method to measure uncertainty in pre-trained prediction models by providing sets of valid class predictions that exceed a given significance level. Here, we first define $d_M$ as the non-conformity measure that assesses the strangeness of an unseen sample. Next, we derive a separate calibration set $S_{calib} = \{(x_1, y_1),..., (x_n, y_n)\}$ with samples that stem from the same distribution as the training data but are unseen to the model. The non-conformity score $d_M$ is computed for every sample in the calibration set. To evaluate how different an unseen sample $x_u$ is from the training distribution, its non-conformity score $d_{M,u}$ is then compared to the non-conformity scores $d_{M,j}$ of the calibration set for both binary 5-year relapse class labels $y_c$, such that

$$p_c(d_{M,u}) = \frac{\left|\left\{j = 1,...,n : y_j = y_c \ and \ d_{M,j} \geq d_{M,u}\right\}\right|}{\left|\left\{j = 1,...,n : y_j = y_c\right\}\right|}$$

where $p_c(d_{M,u})$ refers to the p-value (distinct from the statistical p-value) for a given class $y_c$. High p-values indicate high conformity with the training distribution, since most calibration examples expressed higher non-conformity scores than $x_u$ [33].

The maximum p-value among both 5-year relapse classes is defined as the credibility $Cred_u$ of an unseen sample, such that

$$Cred_u = max_u(p_c(d_{M,u}))$$

This credibility score quantifies how close a given sample is to the model's learned distribution, based on the unseen calibration dataset, and is expected to correlate with the validity of the final risk prediction.

The validation split of those UKEhv sub-domains present in the training set serves as the calibration data when applying the Credibility Estimation setup to PCAI and the baseline model, such that it consists of data from the UKE-first, UKE-second and UKE-scanner datasets for the former and of data from UKE-first only for the latter.

#### Color adaptation

With the aim to enable valid predictions even on images where PCAI shows a low credibility, a color adaptation setup to map the color of those images to the color scheme of the training distribution is established (Fig. S2C, S3, S4A). Color is a strong separator between datasets used in this work. In detail, we propose a cluster-based histogram matching procedure, which Dietrich et al. found to improve over matching randomly to a training domain image (Fig. S3)

[34]. For this, we first derive 8 k-means clusters from the histograms of the training data in the HSV space, using Wasserstein distance as the distance measure. This clustering approach smoothes the effect of outliers while preserving inherent type differences inside the dataset. Using the CE setup described above, we define a threshold on the credibility scores such that 75% of the calibration set (i.e. the validation data of the training domains UKE-first, UKE-second and UKE-scanner) expresses higher credibility scores. During inference of the PCAI model, we then match the histograms of samples of the test set that express credibility scores below the defined threshold with the histogram of the closest cluster in the training data and feed those adapted samples through the deep learning network again (Fig. S4A). The 75% threshold as well as the number of 8 clusters was chosen by optimizing the increase in 5-year AUROC on the validation sets of the internal UKEhv sub-domains. Performance metrics of PCAI reported in this manuscript are calculated on the predictions of the resulting combination of raw and color adapted samples, based on their credibility. Fig. S4B shows histograms and patches of an exemplary sample from the MMX dataset before and after color adaptation.

**Cancer indicator**

To indicate cancer-containing regions, the CI is trained on patch-wise cancer vs non-cancer labels extracted from segmentation masks of the PANDA dataset [22]. Fig. S1 shows an exemplary slide of the PANDA dataset with mask overlay in green for healthy tissue and red for cancerous regions with exemplary patches with a side length of 256 pixels with healthy, cancerous, or rejected labels. A total of 4,459,674 training and 504,027 test set patches were extracted from the dataset. The CI model consists of an CNN-encoder, specifically the Efficientnet-b0 architecture, and a subsequent fully connected classification layer. The CI is trained using patch-wise labels extracted from segmentation annotations provided by expert (uro-) pathologists in the PANDA challenge. With this, it achieves an AUROC of 0.94 on the PANDA test set patches of previously unseen slides. In the overall PCAI model, CI is utilized to reduce noise and redundancy in our risk prediction on biopsies. It is used to predict cancer heatmaps on our biopsy datasets and select the 100 patches with the highest predicted cancer during inference into our PCAI model.

**PCAI risk groups**

To enhance interpretability of our PCAI risk score, we can stratify the patients into risk groups by *k*-means clustering on survival curves by taking the risk $r_i \in R$ for each individual $i$ and perform a 1-dimensional *k*-means clustering algorithm to obtain $k$ distinct groups of patients [35]. To estimate the maximum number of groups that are statistically significant in terms of outcome, a Fleming Harrington-weighted pairwise log-rank test was used on a separate validation set as suggested by Li et al. [36]. P-values for the pairwise logrank test can be found in the appendix in Fig. S5.

### Supplemental Figures & Tables

| | | UKE-first | UKE-second | UKE-scanner | UKE-thin | UKE-thick | UKE-long |
|---|---|---|---|---|---|---|---|
| | patients (images) | 8123 | 7156 | 8114 | 1602 | 1574 | 1667 |
| | age [years], mean ± SD | 63.5 ± 6.1 | 63.6 ± 6.1 | 63.5 ± 6.1 | 63.2 ± 6 | 63.2 ± 5.9 | 63.2 ± 6 |
| | censoring [%] | 61.3 | 60.9 | 61.3 | 67.4 | 67.6 | 67.4 |
| | median survival [years] | 1.6 | 1.6 | 1.6 | 2.4 | 2.4 | 2.4 |
| | median followup [years] | 8 | 8 | 8 | 7.2 | 7.2 | 7.7 |
| ISUP | 0 | 407 (5.01%) | 370 (5.17%) | 405 (4.99%) | 98 (6.12%) | 96 (6.10%) | 103 (6.18%) |
| | 1 | 1792 (22.06%) | 1557 (21.76%) | 1789 (22.05%) | 305 (19.04%) | 304 (19.31%) | 322 (19.32%) |
| | 2 | 4001 (49.26%) | 3512 (49.08%) | 3997 (49.26%) | 879 (54.87%) | 864 (54.89%) | 911 (54.65%) |
| | 3 | 1366 (16.82%) | 1223 (17.09%) | 1366 (16.84%) | 253 (15.79%) | 243 (15.44%) | 262 (15.72%) |
| | 4 | 109 (1.34%) | 94 (1.31%) | 109 (1.34%) | 19 (1.19%) | 19 (1.21%) | 21 (1.26%) |
| | 5 | 448 (5.52%) | 400 (5.59%) | 448 (5.52%) | 48 (3.00%) | 48 (3.05%) | 48 (2.88%) |
| event type | BCR | 3084 (37.97%) | 2745 (38.36%) | 3081 (37.97%) | 518 (32.33%) | 506 (32.15%) | 539 (32.33%) |
| | FU | 4978 (61.28%) | 4355 (60.86%) | 4972 (61.28%) | 1080 (67.42%) | 1064 (67.60%) | 1123 (67.37%) |
| | META | 61 (0.75%) | 56 (0.78%) | 61 (0.75%) | 4 (0.25%) | 4 (0.25%) | 5 (0.30%) |
| T-stage | ≤ T1 | 2 (0.02%) | 2 (0.03%) | 2 (0.02%) | | | |
| | T2 | 4940 (60.81%) | 4301 (60.10%) | 4932 (60.78%) | 976 (60.92%) | 958 (60.86%) | 1021 (61.25%) |
| | T3 | 3120 (38.41%) | 2796 (39.07%) | 3119 (38.44%) | 610 (38.08%) | 601 (38.18%) | 628 (37.67%) |
| | T4 | 61 (0.75%) | 57 (0.80%) | 61 (0.75%) | 16 (1.00%) | 15 (0.95%) | 18 (1.08%) |
| N-stage | N0 | 4290 (86.39%) | 3719 (85.61%) | 4284 (86.37%) | 923 (90.22%) | 907 (90.25%) | 971 (90.49%) |
| | N1 | 676 (13.61%) | 625 (14.39%) | 676 (13.63%) | 100 (9.78%) | 98 (9.75%) | 102 (9.51%) |
| M-stage | M0 | 6306 (78.44%) | 5499 (77.67%) | 6298 (78.43%) | 1260 (78.95%) | 1237 (78.89%) | 1313 (79.05%) |
| | M1 | 1733 (21.56%) | 1581 (22.33%) | 1732 (21.57%) | 336 (21.05%) | 331 (21.11%) | 348 (20.95%) |

**Table S1** Basic patient characteristics of all UKEhv sub-dataset experiments showing number of unique patients (that is the same as the number of images), age, PSA level at RP, censoring rate, median survival and follow-up time in months, the event type classification (BCR=biochemical recurrence, META=metastasis, FU=lost to follow-up), ISUP, pathological T-, N- and M-stage.

| (sub-) dataset | tissue type | #pixels long edge | #pixels short edge | scanner | mag. | µm/pixel |
|---|---|---|---|---|---|---|
| UKE-first | T | 2900 ± 200 | 2900 ± 200 | APE | 40x | 0.25 |
| UKE-second | T | 2900 ± 0 | 2900 ± 0 | APE | 40x | 0.25 |
| UKE-scanner | T | 6100 ± 0 | 6100 ± 0 | 3DH | 80x | 0.125 |
| UKE-thin | T | 2900 ± 0 | 2900 ± 100 | APE | 40x | 0.25 |
| UKE-thick | T | 2900 ± 0 | 2900 ± 0 | APE | 40x | 0.25 |
| UKE-long | T | 2900 ± 0 | 2900 ± 0 | APE | 40x | 0.25 |
| UKE-sealed | T | 3100 ± 200 | 3100 ± 200 | APE | 40x | 0.25 |
| NYU | T | 1800 ± 0 | 1800 ± 0 | APE | 20x | 0.5 |
| JHU | T | 3600 ± 0 | 3600 ± 0 | HAM, VEN | 40x | 0.23 |
| UPP | B | 67100 ± 16300 | 28800 ± 8100 | APE | 40x | 0.25 |
| MMX | B | 64900 ± 22000 | 30200 ± 17400 | HAM, VEN | 40x | 0.23 |
| PANDA | B | 26100 ± 8600 | 15900 ± 8900 | APE, 3DH, HAM | 20x | 0.486 |

**Table S2** Basic image properties of this work's image datasets showing the dataset's tissue type (TMA=T or biopsy=B), mean +- std of the number of pixels on the long and short edge of each image, used scanner vendor (APE=Leica Aperio, 3DH=3DHistech, HAM=Hamamatsu, VEN=Ventana), mag.=maximum magnification level and the resulting physical resolution in µm per pixel.

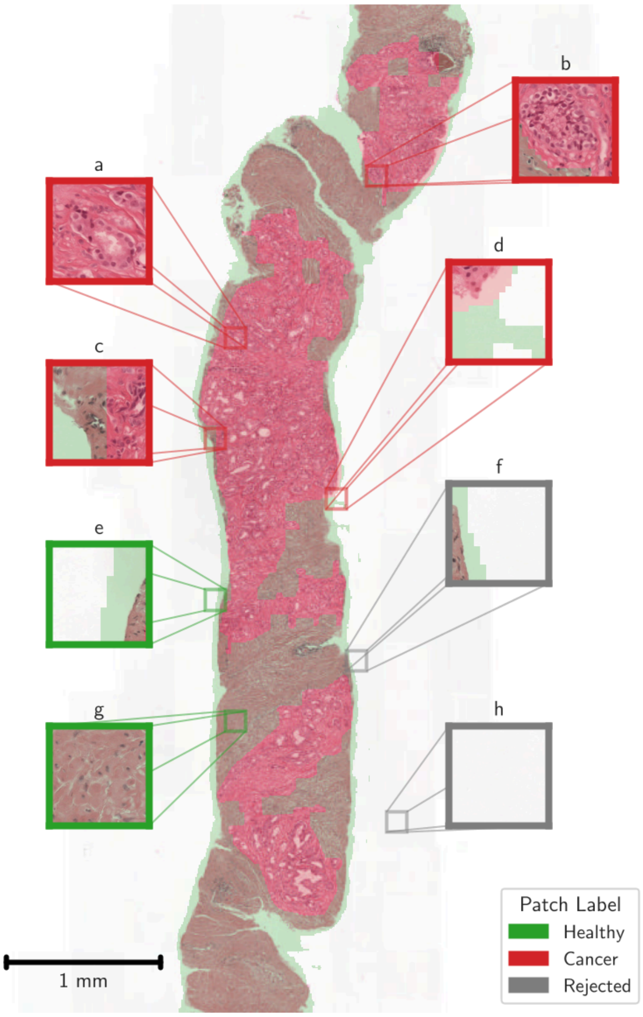


**Figure S1** Exemplary slide with cancer and tissue mask of the PANDA dataset. **A-H** Visualization of potentially extracted patches from the slide. Gray patches are rejected, red patches are labeled as cancerous, and green patches as healthy. Note that the segmentation masks extend not only in the tissue but also in the background in patches in **D-F**.

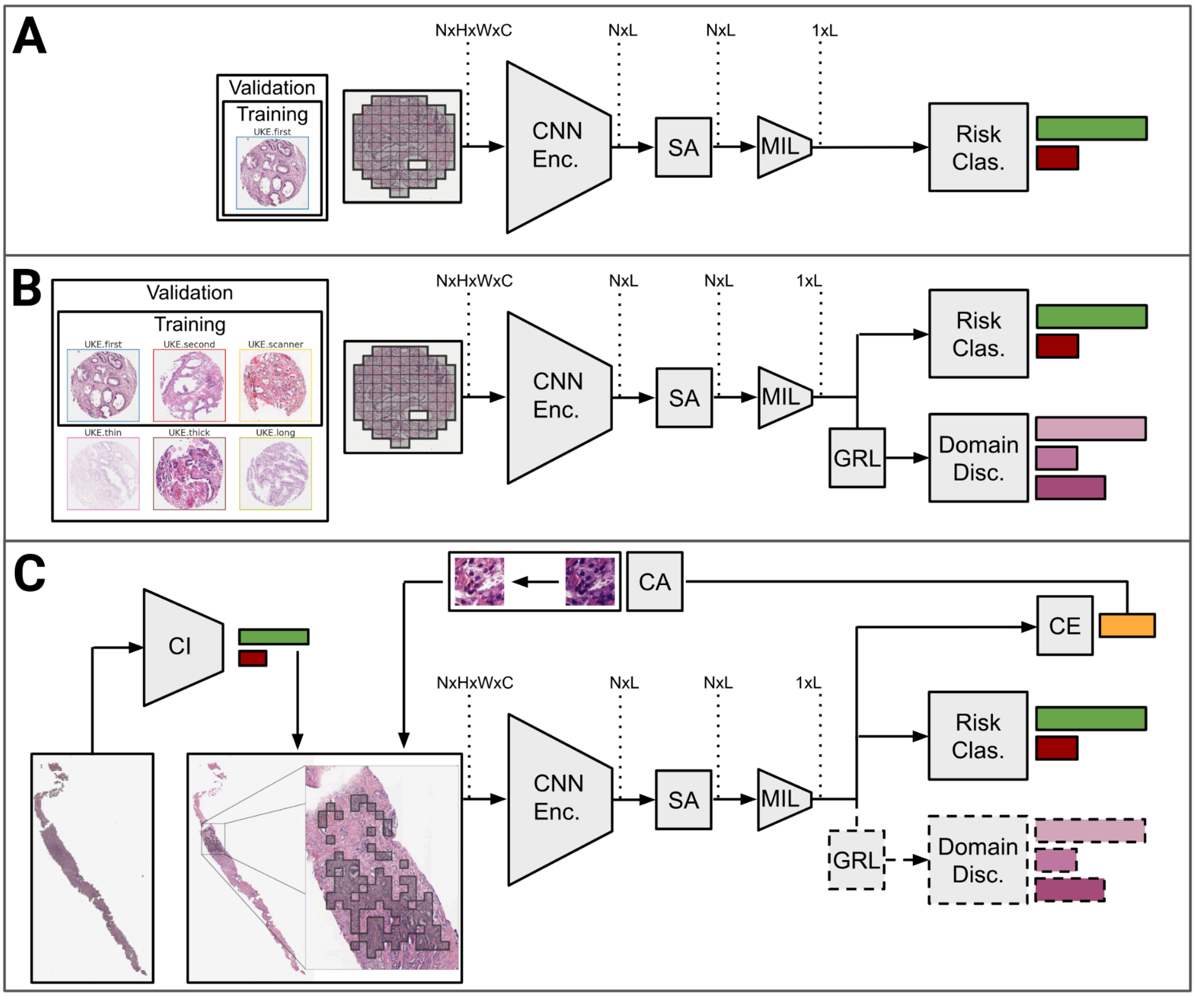


**Figure S2** Architecture and training regime of **A** the BASE risk prediction network, **B** the BASE model including the DA module, and **C** PCAI with added CI based patch sampling, CE, and CA. CNN Enc. = Convolutional neural network encoder; SA = Self-attention layer; MIL= Attention-based multiple instance learning layer; GRL = Gradient reversal layer; Risk Clas. = Risk classifier; Domain Disc. = Domain discriminator; DA = Domain adversarial; CI = Cancer indicator; CE = credibility estimation; CA = color adaptation.

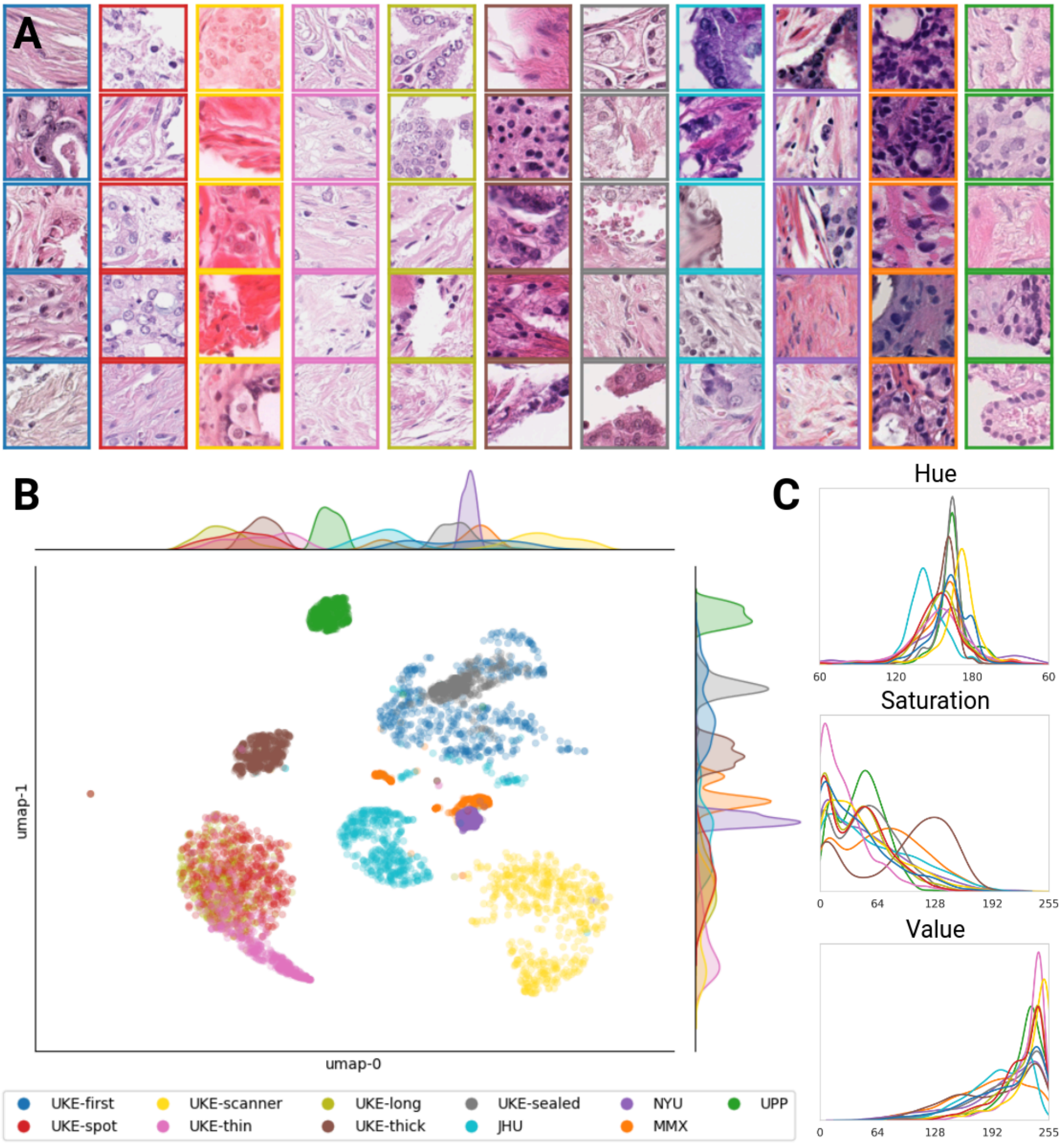


**Figure S3** Overview of color variations across all datasets. **A** Example patches of all datasets used in this study. Color-coded margins depict data origin. **B** UMAP of the HSV histograms. **C** Aggregated hue, saturation, and value histograms of all valid foreground pixels of all images per dataset.

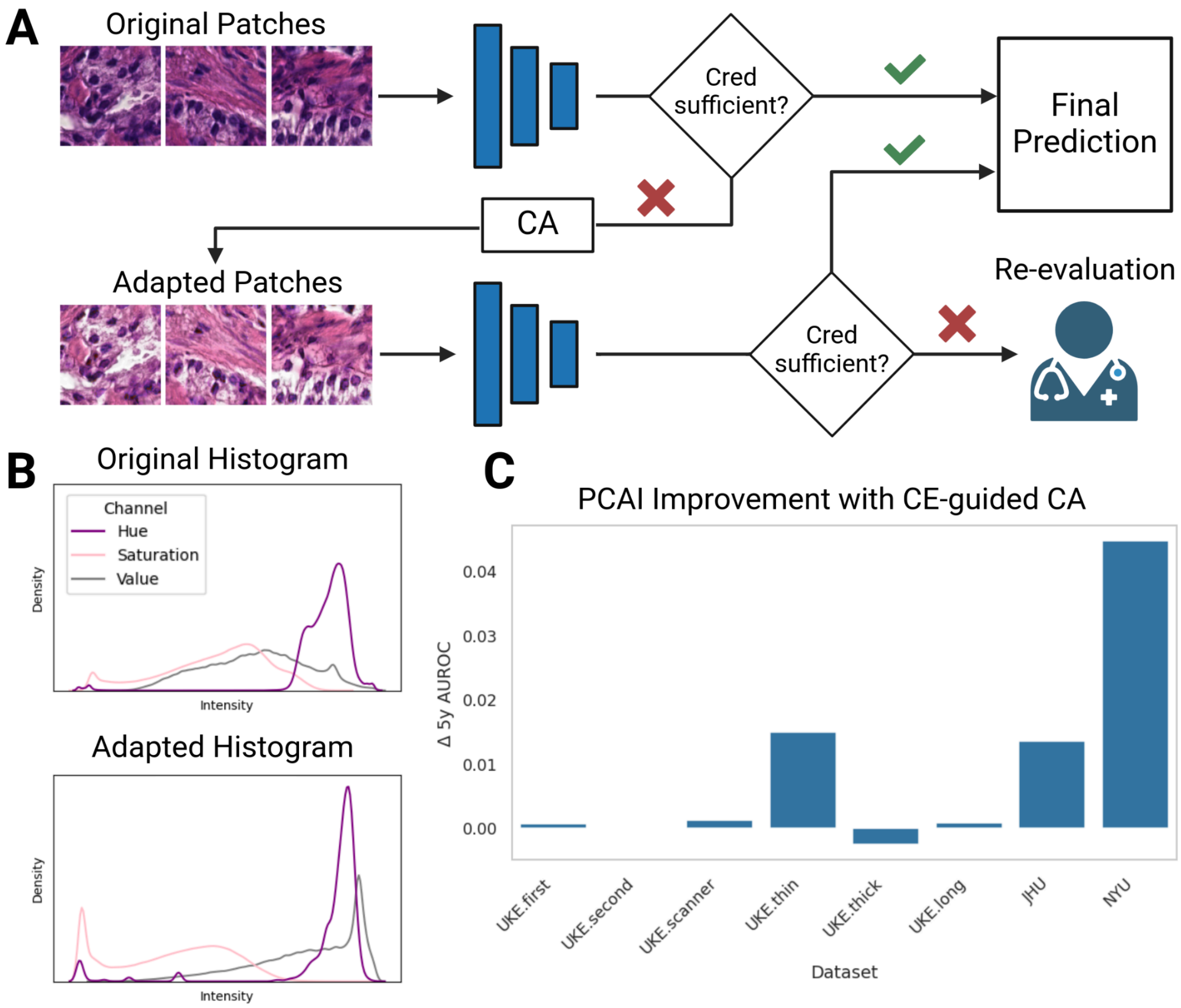


**Figure S4** Credibility-guided color adaptation in PCAI **A** Feedback loop of the credibility-guided color adaptation (CE-CA) procedure. If during initial processing of the image in the deep learning network (blue) sufficient credibility is not reached, the color of the problematic sample is adapted by matching its histogram with the training distribution. If sufficient credibility is still not reached, grading of the images can be conferred to the pathologist . **B** Exemplary HSV histograms of a sample before and after applying CE-CA. **C** Improvement in 5-year AUROC in PCAI when using the proposed CE-CA procedure over predicting on un-altered images only.

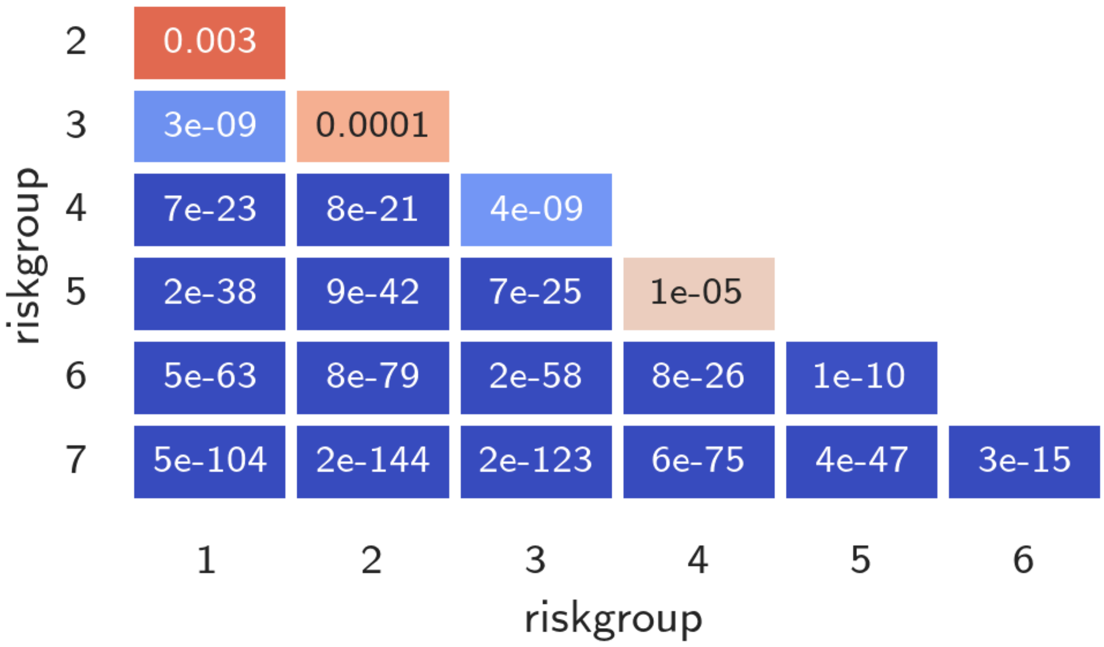


**Figure S5** Results of the pairwise logrank test for the UKEhv datasets based on PCAI predictions. Values below p=0.05 were interpreted as statistically significant.

A

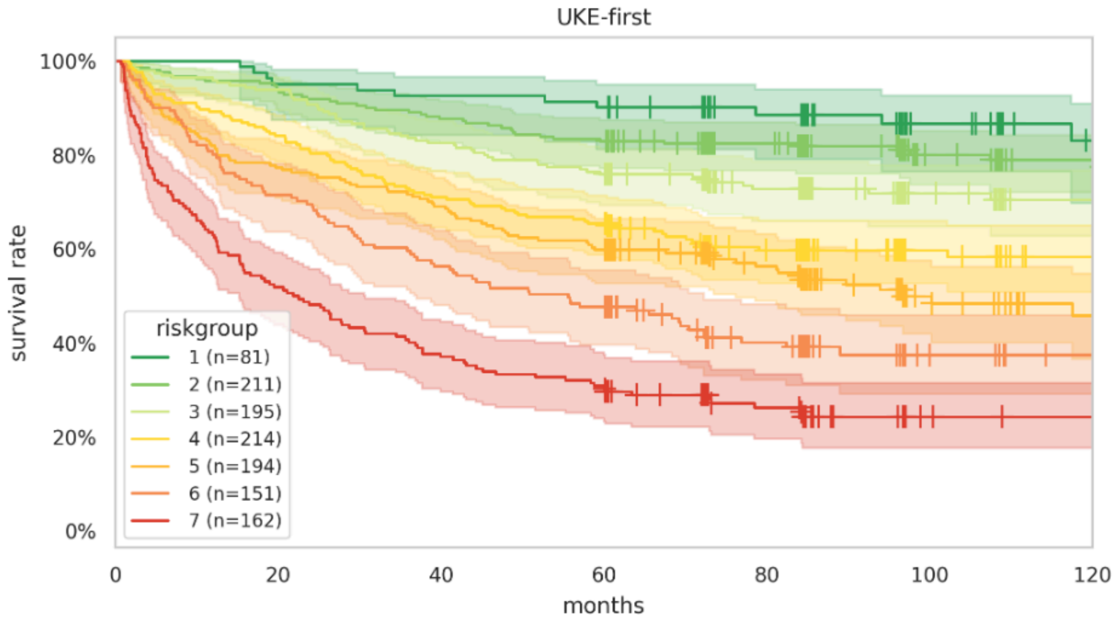


B

| PCAI riskgroup | | time 0 | 20 | 40 | 60 | 80 | 100 | 120 |
|---|---|---|---|---|---|---|---|---|
| | At risk | 81 | 77 | 75 | 73 | 57 | 34 | 22 |
| 1 (n=81) | Censored | 0 | 0 | 0 | 0 | 15 | 37 | 48 |
| | Events | 0 | 4 | 6 | 8 | 9 | 10 | 11 |
| | At risk | 211 | 198 | 185 | 175 | 139 | 73 | 56 |
| 2 (n=211) | Censored | 0 | 0 | 0 | 0 | 35 | 98 | 114 |
| | Events | 0 | 13 | 26 | 36 | 37 | 40 | 41 |
| | At risk | 195 | 183 | 161 | 148 | 110 | 51 | 30 |
| 3 (n=195) | Censored | 0 | 0 | 0 | 0 | 33 | 91 | 111 |
| | Events | 0 | 12 | 34 | 47 | 52 | 53 | 54 |
| | At risk | 214 | 180 | 152 | 140 | 85 | 42 | 28 |
| 4 (n=214) | Censored | 0 | 0 | 0 | 0 | 45 | 88 | 101 |
| | Events | 0 | 34 | 62 | 74 | 84 | 84 | 85 |
| | At risk | 194 | 150 | 134 | 116 | 78 | 32 | 18 |
| 5 (n=194) | Censored | 0 | 0 | 0 | 0 | 33 | 72 | 84 |
| | Events | 0 | 44 | 60 | 78 | 83 | 90 | 92 |
| | At risk | 151 | 108 | 85 | 72 | 41 | 13 | 7 |
| 6 (n=151) | Censored | 0 | 0 | 0 | 0 | 22 | 48 | 54 |
| | Events | 0 | 43 | 66 | 79 | 88 | 90 | 90 |
| | At risk | 162 | 84 | 60 | 50 | 29 | 9 | 7 |
| 7 (n=162) | Censored | 0 | 0 | 0 | 0 | 15 | 33 | 35 |
| | Events | 0 | 78 | 102 | 112 | 118 | 120 | 120 |

**Figure S6** KM curves in **A** with the corresponding at-risk table in **B** for the UKEhv test dataset to visualize the discriminative performance of the PCAI risk grouping.

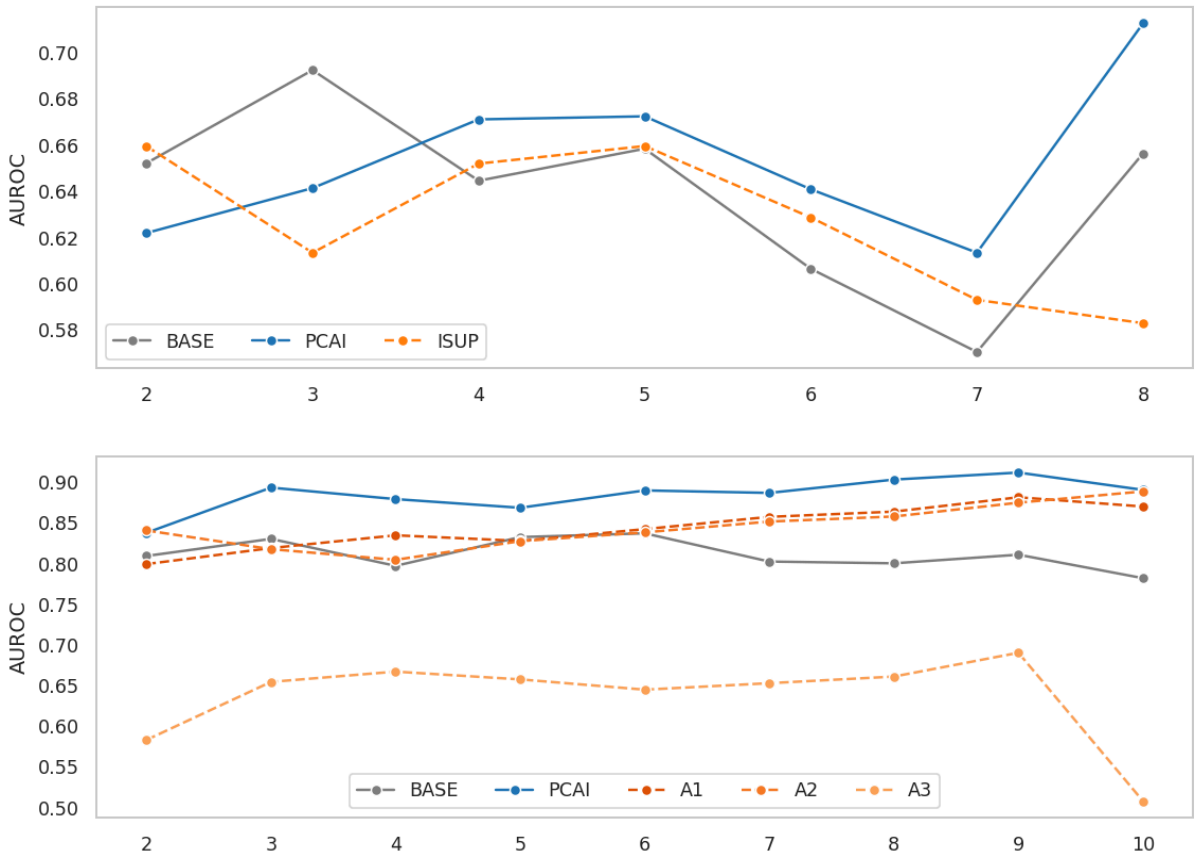


**Figure S7** BASE and PCAI performance compared to human annotators for the UPP and MMX biopsy datasets. The 2-10 year AUROC is shown for each prediction (gray for BASE, blue for PCAI) and human annotation (orange shades). It is interesting to observe that for almost every temporal (year) threshold the PCAI model performs best.
